# Evaluation of a Rapid and Accessible RT-qPCR Approach for SARS-CoV-2 Variant of Concern Identification

**DOI:** 10.1101/2022.02.07.22270629

**Authors:** Priscilla S.-W. Yeung, Hannah Wang, Mamdouh Sibai, Daniel Solis, Fumiko Yamamoto, Naomi Iwai, Becky Jiang, Nathan Hammond, Bernadette Truong, Selamawit Bihon, Suzette Santos, Marilyn Mar, Claire Mai, Kenji O. Mfuh, Jacob A. Miller, ChunHong Huang, Malaya K. Sahoo, James L. Zehnder, Benjamin A. Pinsky

## Abstract

The ability to distinguish between SARS-CoV-2 variants of concern (VOCs) is of ongoing interest due to differences in transmissibility, response to vaccination, clinical prognosis, and therapy. Although detailed genetic characterization requires whole-genome sequencing (WGS), targeted nucleic acid amplification tests can serve a complementary role in clinical settings, as they are more rapid and accessible than sequencing in most laboratories.

We designed and analytically validated a two-reaction multiplex reverse transcription quantitative PCR (RT-qPCR) assay targeting spike protein mutations L452R, E484K, and N501Y in Reaction 1, and del69-70, K417N, and T478K in Reaction 2. This assay had 95-100% agreement with WGS in 502 upper respiratory swabs collected between April 26 and August 1, 2021, consisting of 43 Alpha, 2 Beta, 20 Gamma, 378 Delta, and 59 non-VOC infections. Validation in a separate group of 230 WGS-confirmed Omicron variant samples collected in December 2021 and January 2022 demonstrated 100% agreement.

This RT-qPCR-based approach can be implemented in clinical laboratories already performing SARS-CoV-2 nucleic acid amplification tests to assist in local epidemiological surveillance and clinical decision-making.

## INTRODUCTION

Since the original strain of SARS-CoV-2 virus was first discovered in late 2019, numerous new variants have been identified, including variants of concern (VOCs) Alpha (B.1.1.7 and sublineages), Beta (B.1.351), Gamma (P.1 and sublineages), Delta (B.1.617.2 and sublineages) and Omicron (B.1.1.529 and sublineages) (1). Importantly, these VOCs differ in their clinical prognosis, transmissibility, antibody susceptibility, and response to vaccination (2-22). Whole-genome sequencing (WGS) has played a critical role in identifying the emergence of these new variants (23-25), and millions of distinct sequences have been deposited into public repositories such as the Global Initiative on Sharing Avian Influenza Data consortium’s database (GISAID) (26). However, WGS has a relatively long turnaround time, is labor-intensive, and requires instruments, bioinformatics support, and specially-trained staff that may not be widely available to many clinical laboratories. Therefore, the development of reverse transcription quantitative PCR (RT-qPCR) assays to presumptively type SARS-CoV-2 variants may be an important real-time complement to WGS epidemiologic surveillance, and may directly impact the clinical care of individual patients by informing selection of expensive and potentially difficult-to-source monoclonal antibody therapies (2, 7, 13-17, 20, 21, 27). It is important to note that such presumptive typing assays may provide atypical results in emerging strains due to mutations within primer and/or probe binding sites. As such, they must be intelligently-designed, thoroughly validated, and interpreted carefully.

In this study, we report the design of a multiplex RT-qPCR assay that detects the del69-70, K417N, and T478K mutations in SARS-CoV-2 spike protein and targets the wild-type 69-70 sequence as an internal control. We further evaluate the performance of this assay in combination with our previously described RT-qPCR assay for the detection of L452R, E484K, and N501Y (28), and demonstrate the utility of this targeted mutational analysis to accurately distinguish among VOCs.

## MATERIALS AND METHODS

### Assay Design

The spike protein mutations associated with each variant that are interrogated by the RT-qPCR assays are summarized in Figure 1. In the first reaction (Reaction 1), we utilized our previously described RT-qPCR assay to detect L452R, E484K, and N501Y mutations in spike Receptor Binding Domain (RBD) (28). The present study describes the combination of this assay with a second, newly designed reaction (Reaction 2), which detects the deletion of amino acids 69-70 in the spike N-Terminal Domain (del69-70), as well as K417N and T478K mutations in the RBD. We use allele-specific RT-qPCR with probe sequences designed to maximize the difference in annealing temperature between mutant and wild-type sequences, allowing for differential binding and amplification. The primer/probe sequences for each mutation site are summarized in Table 1, and the guidance for interpretation and reporting is described in Table 2. Additional details are provided in the Supplemental Methods, ssDNA sequences for analytical experiments (Supplemental Table 1), analytical validation data (Supplemental Table 2), and in-silico analysis of primer and probe sequences (Supplemental Figure 1).

**Figure 1.**
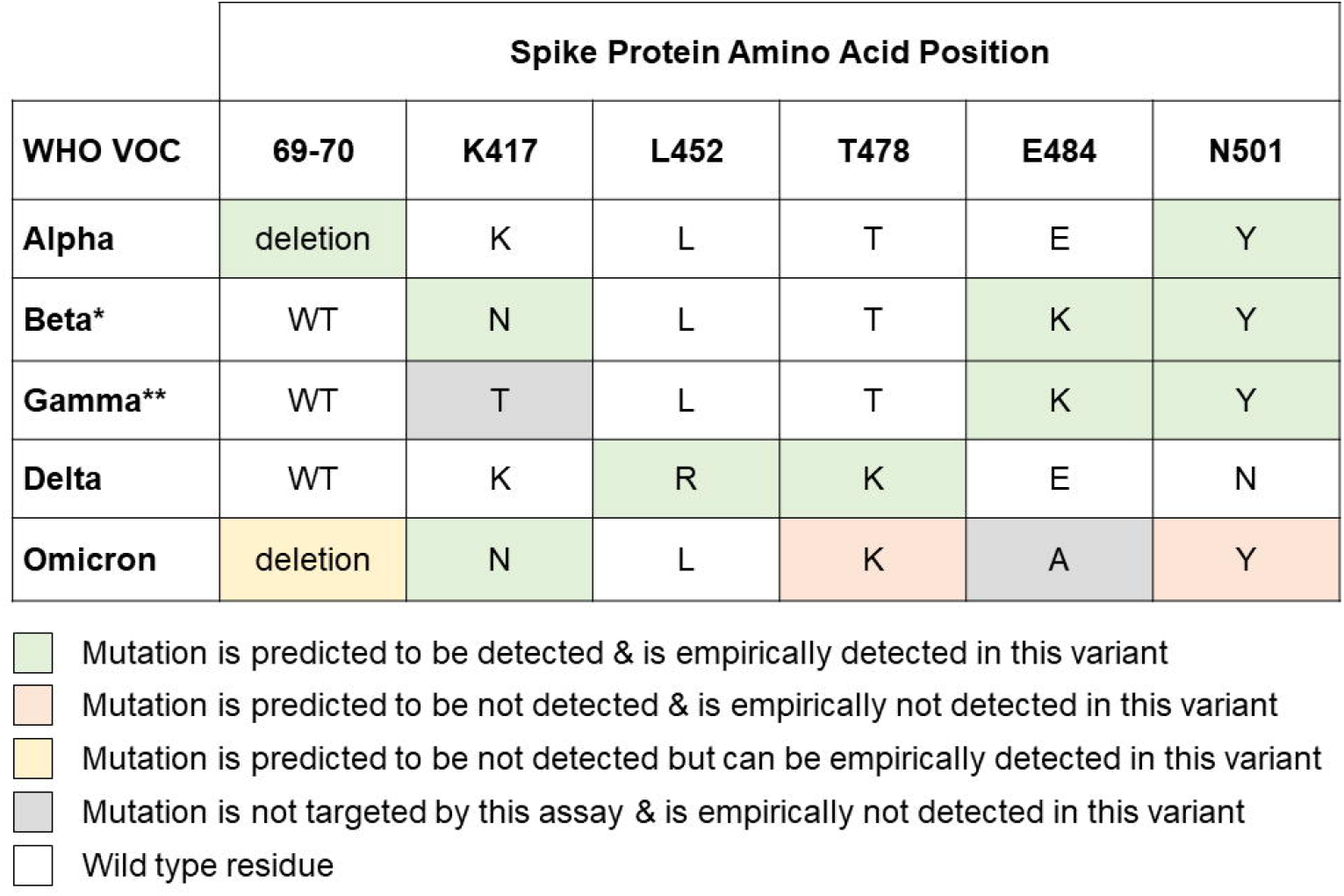
Summary of current World Health Organization (WHO)-designated variants of concern (VOC) along with their expected spike mutations at sites targeted by this two-reaction multiplex SARS-CoV-2 RT-qPCR genotyping approach. These reactions are designed to detect the following mutations: del69-70, K417N, L452R, T478K, E484K, and N501Y. Shading indicates predicted versus empiric performance of this assay for the detection and differentiation of these VOCs. The predicted detection for the VOC Omicron is based on both the sequence at the target sites and known adjacent mutations in the probe binding site. A single asterisk and double asterisk respectively denote a known limitations of the assay in differentiating VOCs Beta and Gamma respectively from the variant of interest Mu.

**Table 1.**
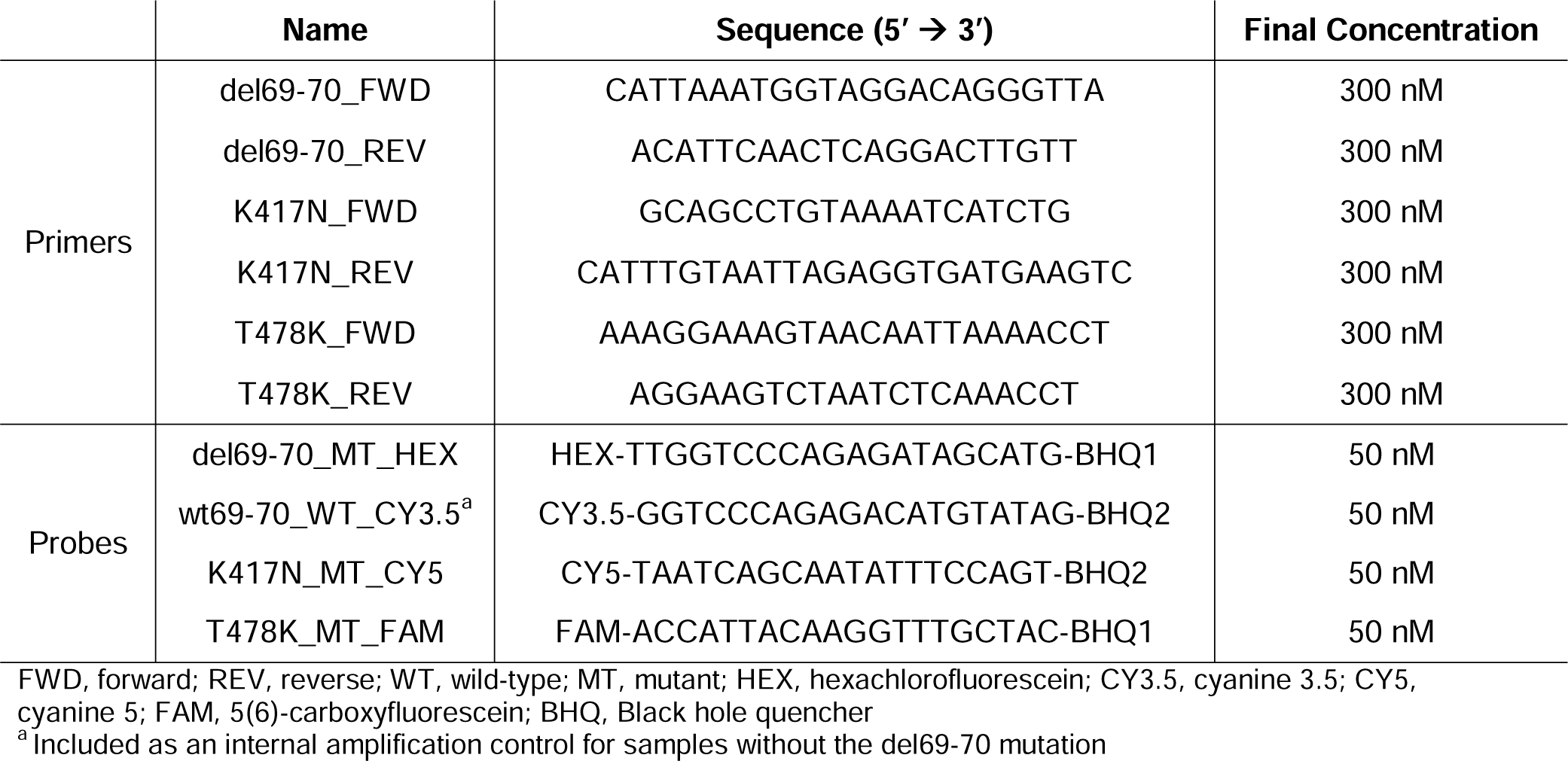
Reaction 2 Primer and Probe Oligonucleotide Sequences

**Table 2.** Interpretation and Reporting of the Two-Reaction Multiplex Genotyping RT-qPCR

| Reaction 1 Results |  |  |  | Reaction 2 Results |  |  |  | Action |
| --- | --- | --- | --- | --- | --- | --- | --- | --- |
| N501WT<br>(ROX/Cy3.5) | N501Y<br>(FAM) | E484K<br>(CY5) | L452R<br>(HEX) | 69_70WT<br>(ROX/Cy3.5) | T478K<br>(FAM) | K417N<br>(CY5) | Del69_70<br>(HEX) | See Interp<br>Comments<sup>a</sup> |
| DTD ≤ 40 | NDET | NDET | DTD ≤ 40 | DTD ≤ 40 | DTD ≤ 40 | DTD ≤ 40 | NDET | Report as 1 |
| DTD ≤ 40 | NDET | NDET | DTD ≤ 40 | DTD ≤ 40 | DTD ≤ 40 | NDET | NDET | Report as 1 |
| DTD ≤ 40 | NDET | NDET | DTD ≤ 40 | NDET | DTD ≤ 40 | DTD ≤ 40 | NDET | Report as 1 |
| NDET | DTD ≤ 40 | DTD ≤ 40 | NDET | DTD ≤ 40 | NDET | DTD ≤ 40 | NDET | Report as 2 |
| NDET | DTD ≤ 40 | DTD ≤ 40 | NDET | DTD ≤ 40 | NDET | NDET | NDET | Report as 3 |
| NDET | DTD ≤ 40 | DTD ≤ 40 | NDET | NDET | NDET | NDET | DTD ≤ 40 | Report as 4 |
| NDET | DTD ≤ 40 | NDET | NDET | NDET | NDET | NDET | DTD ≤ 40 | Report as 4 |
| NDET | NDET | NDET | NDET | NDET | NDET | DTD ≤ 40 | DTD ≤ 40 | Report as 5 |
| NDET | NDET | NDET | NDET | NDET | NDET | DTD ≤ 40 | NDET | Report as 5 |
| NDET | NDET | NDET | NDET | NDET | NDET | NDET | NDET | Unable<br>(Refer to 6) |
| Any target with Cq > 40 or abnormal/inconclusive amplification curves. |  |  |  |  |  |  |  | Review<br>(Refer to 7) |
| Any scenarios not designated above. |  |  |  |  |  |  |  | Review<br>(Refer to 7) |

<sup>a</sup> Result Interpretation Comments
1. Probable Variant of Concern Delta; Increased transmissibility; Decreased susceptibility to BAM\* 2. Possible Variant of Concern Beta versus Variant of Interest Mu; Decreased susceptibility to BAM, ETE, BAM/ETE, CAS, REG\* 3. Possible Variant of Concern Gamma versus Variant of Interest Mu; Decreased susceptibility to BAM, ETE, BAM/ETE, CAS, REG\* 4. Probable Variant of Concern Alpha; Decreased susceptibility to ETE\* 5. Probable Variant of Concern Omicron; Increased transmissibility; Decreased susceptibility to BAM, ETE, BAM/ETE, CAS, IMD, CAS/IMD, CIL, TIX, CIL/TIX, REG; Decreased susceptibility to convalescent plasma 6. Unable to interpret results due to low level of viral RNA 7. **TRIAGE FOR MEDICAL DIRECTOR REVIEW** (Mutations uncommonly seen together, which could indicate possible mixed infection, contamination, or nonspecific amplification. Analyze additional information including previous results, curve shape, etc. Potential new variant or novel mutation in primer/probe sites causing dropout. Consider sequencing if level of viral RNA is sufficient.)
\*Interpretations about mAb susceptibility and plasma susceptibility were based on data from the Stanford Coronavirus Antiviral and Resistance Database (covdb.stanford.edu). Decreased susceptibility to mAb and convalescent plasma defined as >10 fold reduction in neutralization. Monoclonal antibody (mAb) abbreviations: **BAM**: Bamlanivimab/LY-CoV555, **CAS**: Casirivimab/REGN10933, **IMD**: Imdevimab/REGN10987, **CAS/IMD**: Casirivimab+imdevimab/REGN-COV2, **ETE**: Etesevimab/LY-CoV016/JS016/CB6, **CIL**: Cilgavimab/COV2-2130/AZD1061, **TIX**: Tixagevimab/COV2-2196/AZD8895, **TIX/CIL**: Tixagevimab+Cilgavimab, **BAM/ETE**: Bamlanivimab+Etesevimab, **SOT**: Sotrovimab/Vir-7831/S309, **REG**: Regdanvimab/CT-P59.

### Clinical Specimens

The samples included in the initial phase of this study were upper respiratory swab specimens collected from patients as part of routine clinical care from April 26, 2021 to August 1, 2021. Testing was performed at Stanford Clinical Virology Laboratory, which provides virologic testing for all Stanford-affiliated hospitals and outpatient centers in the San Francisco Bay Area. These initial SARS-CoV-2 nucleic acid amplification tests (NAATs) tests prior to genotyping were conducted according to manufacturer and emergency authorization instructions as previously described (28), and in the Supplemental Methods. All samples that tested positive for SARS-CoV-2 RNA were reflexed to genotyping. We then excluded samples that were initially tested by laboratory-based methods with cycle threshold values (Ct) ≥35 or relative light units (RLU) ≤1100. We included all available samples initially tested at or near the point of care as Ct data was not readily available for real-time specimen triage for these samples. We also excluded follow-up specimens to eliminate patient-level duplicates. Subsequent validation of this assay for Omicron variant detection was conducted using a convenience set of 230 Omicron variant samples with available WGS data collected between December 2, 2021 and January 5, 2022. This study was conducted with Stanford institutional review board approval (protocol 57519), and individual consent was waived.

### Whole-Genome Sequencing

To validate the genotyping RT-qPCR reactions, we tested their performance against WGS in a subset of the samples in the initial April 26, 2021 to August 1, 2021 cohort with Ct <30. Samples with non-dominant variant typing by RT-qPCR were prioritized for sequencing, with the remaining isolates chosen randomly to fill a sequencing run. WGS was conducted as described previously, using a lab-developed pipeline consisting of long-range PCR, followed by fragmentation, then single-end 150-cycle sequencing using MiSeq reagent kit V3 (Illumina, San Diego, CA) (28). Genomes were assembled via a custom assembly and bioinformatics pipeline using NCBI NC_045512.2 as reference. Whole-genome sequences with ≥75% genome coverage to a depth of at least 10 reads were accepted for interpretation. Mutation calling required a depth of at least 12 reads with a minimum variant frequency of 20%. PANGO lineage assignment was performed using https://pangolin.cog-uk.io/ running pangolin version 3.1.17, while Nextclade Web v1.13.1 and auspice.us 0.8.0. were used to perform phylogenetic placement (3, 29, 30). Both lineage and clade assignments were performed on February 1, 2022. WGS data was deposited in GISAID (Supplemental Table 3).

### Statistical Analysis

Positive percent agreement (PPA) and negative percent agreement (NPA) were reported with Clopper-Pearson score 95% binomial confidence intervals (CI) using WGS as the reference method. Analyses were conducted using the R statistical software package. This study was reported in accordance with Standards for the Reporting of Diagnostic Accuracy Studies (STARD) guidelines.

## RESULTS

During the initial study period of April 26, 2021 to August 1, 2021, the Stanford Clinical Virology Laboratory received 102,158 specimens from 70,544 unique individuals. A total of 1,657 samples from unique individuals tested positive for SARS-CoV-2, of which 1,093 (66%) had genotyping RT-qPCR Reaction 1 and Reaction 2 performed, and 502 (30.3%) had successful WGS performed (Supplemental Figure 2). The lower limit of detection for the mutation site probes in Reaction 2 (in copies per µL template) were 14.8 for del69-70 (HEX), 16.4 for K417N (CY5), and 2.1 for T478K (FAM). Median number of aligned reads for WGS was 485,870 (interquartile range [IQR] 289,363-655,481), while median genome coverage to a depth of at least 10 reads was 99.3% (IQR 97.1-99.3%). Of note, Reaction 1 was performed in near real-time, while Reaction 2 was performed retrospectively. Overall, this subset of sequenced samples, had patient and testing characteristics that closely resembled those of the larger cohorts (Supplemental Table 4).

The assay resulted as “Unable to Genotype” in 152 of 1,093 samples (14%) due to lack of amplification of any target in either or both reactions. Assay failure occurred predominantly in samples originally tested at or near the point of care (119/341, 35%), where all positive samples were triaged for genotyping without any filter. In contrast, assay failure occurred much less frequently in samples originally tested in the moderate-to-high complexity virology lab (33/752, 4%), where samples with lower viral RNA levels (Ct≥35) were not triaged for genotyping. In the latter group of 752 samples tested in the virology lab, 601 had known Ct values. Among these 601 samples, 68 had Ct > 30, of which 11/68 (16%) failed amplification.

PPA and NPA for the six individual mutations targeted by the genotyping RT-qPCRs as compared to WGS were calculated from the 502 samples with both RT-qPCR and WGS performed (Supplemental Figure 2). The number of samples positive for each mutation reflects the natural prevalence of each of these mutations during this time period. For the combination of Reactions 1 and 2, the PPAs for del69-70, L452R, T478K, E484K, and N501Y were 100% (Table 3). Across all six loci, only K417N had a false negative, resulting in a PPA of 96% (27/28). In this sample, WGS showed a synonymous T to C mutation at position 1254 of the spike gene corresponding to amino acid position 418, changing the codon from ATT to ATC. This single base pair substitution likely decreased the annealing temperature, causing probe dropout and a false negative result.

**Table 3.** Comparison of RT-qPCR and WGS Results for SARS-CoV-2 Spike Gene Mutation Detection in the Initial Cohort (n=502)

| Spike Mutations |  | WGS pos | WGS neg | PPA (95% CI) | NPA (95% CI) |
| --- | --- | --- | --- | --- | --- |
| **Del69-70** | **RT-qPCR pos** | 43 | 0 | 100% (92-100%) | 100% (99-100%) |
|  | **RT-qPCR neg** | 0 | 459 |  |  |
| **K417N** | **RT-qPCR pos** | 27 | 0 | 96% (82-100%) | 100% (99-100%) |
|  | **RT-qPCR neg** | 1<sup>a</sup> | 474 |  |  |
| **L452R** | **RT-qPCR pos** | 403 | 5<sup>b</sup> | 100% (99-100%) | 95% (89-98%) |
|  | **RT-qPCR neg** | 0 | 94 |  |  |
| **T478K** | **RT-qPCR pos** | 379 | 0 | 100% (99-100%) | 100% (97-100%) |
|  | **RT-qPCR neg** | 0 | 123 |  |  |
| **E484K** | **RT-qPCR pos** | 35 | 3<sup>c</sup> | 100% (90-100%) | 99% (98-100%) |
|  | **RT-qPCR neg** | 0 | 464 |  |  |
| **N501Y** | **RT-qPCR pos** | 70 | 0 | 100% (95-100%) | 100% (99-100%) |
|  | **RT-qPCR neg** | 0 | 432 |  |  |

RT-qPCR, reverse transcription quantitative polymerase chain reaction; WGS, whole-genome sequencing; PPA, positive percent agreement; NPA, negative percent agreement; CI, confidence interval
<sup>a</sup> False negative RT-qPCR result due to synonymous mutation in spike gene amino acid position 418 (codon ATT -> ATC) causing probe dropout.
<sup>b</sup> False negative WGS results due to insufficient read count (<12) at this codon. Manual review of sequences revealed 3-9 mutant reads in each sample.
<sup>c</sup> These three samples were found on WGS to be positive for E484Q. While positive for the E484K target on RT-qPCR, these samples had a distinct blunted amplification curve associated with E484Q as previously described (31).

The NPAs for del69-70, K417N, T478K, and N501Y were 100% (Table 3). L452R had an NPA of 95% (94/99) and E484K had an NPA of 99% (464/467). At the L452 locus, there were five samples positive for L452R mutation by RT-qPCR that were negative by WGS. Manual review of the WGS data showed that these were likely false negative WGS results due to insufficient (<12 reads) coverage at this codon. There were 3-9 reads containing the L452R mutation identified in the WGS primary data in each of these five samples. These five samples were all in the Delta lineage based on mutations found at other positions by sequencing.

For the E484K target, there were three samples that tested positive for the E to K mutation but in fact had a E484Q mutation determined by WGS. In both the E to K mutation (GAA to AAA) as well as the E to Q mutation (GAA to CAA), there was a single base substitution at the first position of the codon resulting in nonspecific probe binding. These three samples had a distinct blunted amplification curve with high Ct values associated with E484Q, as previously described (31). While this cross-reactivity is a limitation of the E484K probe design, the unusually-shaped amplification curves were identified and flagged for medical director review as part of assay interpretation protocol (Table 2). Presumptive typing for such cases would need to be deferred until WGS confirmation.

Of note, there was a subset of variant AY.2, involving four specimens in our cohort, that had a V70F mutation causing both del69-70 and wt69-70 probes not to bind. However, because this variant would have T478K and K417N detected, the wt69-70 signal was not needed as an amplification control. This subset of the AY.2 genotype is expected to be positive for L452R and N501 wildtype internal control while negative for N501Y and E484K in Reaction 1, and positive for K417N and T4748K while negative for del69-70 and wildtype 69-70 internal control in Reaction 2. This scenario has been reflected in the clinical interpretation table (Table 2).

SARS-CoV-2 positive specimens collected starting December 2, 2021 began to show an unusual combination of mutations: presence of K417N and del69-70 only in Reaction 2, with all targets including internal control N501 not detected in Reaction 1. Based on in-silico analysis, we determined that these cases likely represented the Omicron variant. While most Omicron variant strains possess del69-70, K417N, T478K, and N501Y mutations, they also have mutations at A67V, S477N, and Q498R, which would be predicted to interfere with binding of the del69-70/wt69-70, T478K, and N501Y/N501 probes, respectively. Although the E484K probe demonstrated cross-reactivity with strains containing the E484Q mutation, as described above, the E484K probe did not detect the E484A mutation in the Omicron variant since it differed by two or more bases. The del69-70 probe likely was able to retain some degree of binding due to the wider melting temperature differential of a 6-nucleotide deletion compared to a point mutation. As such, we validated this assay for Omicron detection using a set of 230 SARS-CoV-2 positive samples confirmed to be Omicron by WGS. We found that the unique pattern of K417N and del69-70 in Reaction 2, along with failure to amplify any target including internal control in Reaction 1, was present in 230/230 (100%, 95% CI 98-100%) Omicron samples tested. This pattern was not seen in any of the 1,093 non-Omicron samples previously genotyped.

We next predicted the WHO variant designation of samples using RT-qPCR and correlated them with the PANGO lineage assignments based on WGS data (Table 4). Mapping the genotyping results of the cohort based on RT-qPCR mutation analysis onto the Nextclade phylogenetic tree demonstrated close correlation with their WHO variant designations (Figure 2). Among the 732 clinical samples that were tested by both RT-qPCR and WGS, 43 samples (5.9%) were Alpha (B.1.1.7 or Q.3), 2 samples (0.3%) were Beta (B.1.351), 20 samples (2.7%) were Gamma (P.1 and sublineages), 378 samples (51.6%) were Delta (B.1.617.2 or AY sublineages), and 230 samples (31.4%) were Omicron (B.1.1.529 or BA sublineages). There were no RT-qPCR false negatives in assigning samples to these lineages. In addition, there were 59 samples (8.1%) tested by WGS that did not correspond to a WHO VOC as of February 2, 2022. Within this subset, there were 4 samples that were erroneously assigned as Gamma and 1 that was assigned as Beta by RT-qPCR. By WGS, these samples were variant of interest (VOI) Mu (B.1.621 or BB.2). This variant shares mutations E484K and N501Y with both the Beta and Gamma variants. A subset of Mu also includes the K417N mutation which is seen in the Beta variant. Thus, our PCR assay could not distinguish VOI Mu from VOCs Beta and Gamma. Our interpretation table information reflects this limitation (Table 2). The remaining 54 samples did not contain mutation patterns associated with VOCs by either RT-qPCR or WGS.

**Table 4.** Comparison of RT-qPCR and WGS for SARS-CoV-2 Variant of Concern Detection (n=732)

| WHO VOC | WGS | RT-qPCR |  |  |  |  |  |  |
| --- | --- | --- | --- | --- | --- | --- | --- | --- |
|  | PANGO lineage | Alpha | Beta | Gamma | Delta | Omicron | Not a VOC | All |
| Alpha | All Alpha | 43 | - | - | - | - | - | 43 |
|  | B.1.1.7 | 37 | - | - | - | - | - | 37 |
|  | Q.3 | 6 | - | - | - | - | - | 6 |
| Beta | Beta | - | 2 | - | - | - | - | 2 |
|  | B.1.351 | - | 2 | - | - | - | - | 2 |
| Gamma | All Gamma | - | - | 20 | - | - | - | 20 |
|  | P.1 | - | - | 13 | - | - | - | 13 |
|  | P.1.10 | - | - | 5 | - | - | - | 5 |
|  | P.1.17 | - | - | 2 | - | - | - | 2 |
| Delta | All Delta | - | - | - | 378 | - | - | 378 |
|  | B.1.617.2 | - | - | - | 29 | - | - | 29 |
|  | AY.1 | - | - | - | 20 | - | - | 20 |
|  | AY.2 | - | - | - | 5 | - | - | 5 |
|  | AY.3 | - | - | - | 5 | - | - | 5 |
|  | AY.4 | - | - | - | 1 | - | - | 1 |
|  | AY.13 | - | - | - | 32 | - | - | 32 |
|  | AY.14 | - | - | - | 59 | - | - | 59 |
|  | AY.19 | - | - | - | 1 | - | - | 1 |
|  | AY.20 | - | - | - | 5 | - | - | 5 |
|  | AY.23 | - | - | - | 1 | - | - | 1 |
|  | AY.25 | - | - | - | 7 | - | - | 7 |
|  | AY.25.1 | - | - | - | 25 | - | - | 25 |
|  | AY.26 | - | - | - | 15 | - | - | 15 |
|  | AY.35 | - | - | - | 2 | - | - | 2 |
|  | AY.43 | - | - | - | 2 | - | - | 2 |
|  | AY.44 | - | - | - | 77 | - | - | 77 |
|  | AY.46.2 | - | - | - | 1 | - | - | 1 |
|  | AY.47 | - | - | - | 8 | - | - | 8 |
|  | AY.48 | - | - | - | 1 | - | - | 1 |
|  | AY.52 | - | - | - | 1 | - | - | 1 |
|  | AY.54 | - | - | - | 3 | - | - | 3 |
|  | AY.59 | - | - | - | 1 | - | - | 1 |
|  | AY.62 | - | - | - | 1 | - | - | 1 |
|  | AY.67 | - | - | - | 3 | - | - | 3 |
|  | AY.74 | - | - | - | 1 | - | - | 1 |
|  | AY.75 | - | - | - | 10 | - | - | 10 |
|  | AY.98.1 | - | - | - | 1 | - | - | 1 |
|  | AY.100 | - | - | - | 3 | - | - | 3 |
|  | AY.103 | - | - | - | 26 | - | - | 26 |
|  | AY.110 | - | - | - | 9 | - | - | 9 |
|  | AY.114 | - | - | - | 1 | - | - | 1 |
|  | AY.116.1 | - | - | - | 2 | - | - | 2 |
|  | AY.118 | - | - | - | 5 | - | - | 5 |
|  | AY.119 | - | - | - | 4 | - | - | 4 |
|  | AY.120.1 | - | - | - | 1 | - | - | 1 |
|  | AY.121 | - | - | - | 3 | - | - | 3 |
|  | AY.122 | - | - | - | 5 | - | - | 5 |
|  | AY.126 | - | - | - | 2 | - | - | 2 |
| Omicron | All Omicron | - | - | - | - | 230 | - | 230 |
|  | B.1.1.529/BA.1 | - | - | - | - | 123 | - | 123 |
|  | BA.1.1 | - | - | - | - | 107 | - | 107 |
| **Not a VOC** | All Non- VOC | - | 1 | 4 | - | - | 54 | 59 |
|  | A.2.5 | - | - | - | - | - | 6 | 6 |
|  | B.1 | - | - | - | - | - | 3 | 3 |
|  | B.1.1.318 | - | - | - | - | - | 1 | 1 |
|  | B.1.1.519 | - | - | - | - | - | 1 | 1 |
|  | B.1.311 | - | - | - | - | - | 1 | 1 |
|  | B.1.427 | - | - | - | - | - | 3 | 3 |
|  | B.1.429 | - | - | - | - | - | 8 | 8 |
|  | B.1.526 | - | - | - | - | - | 10 | 10 |
|  | B.1.621<sup>a</sup> | - | 1 | 2 | - | - | - | 3 |
|  | BB.2<sup>a</sup> | - | - | 2 | - | - | - | 2 |
|  | B.1.627 | - | - | - | - | - | 1 | 1 |
|  | B.1.637 | - | - | - | - | - | 11 | 11 |
|  | XB | - | - | - | - | - | 9 | 9 |
| **All** | All Variants | 43 | 3 | 24 | 378 | 230 | 54 | 732 |

WGS, whole-genome sequencing; RT-qPCR, reverse transcription quantitative polymerase chain reaction; WHO, World Health Organization; VOC, variant of concern
<sup>a</sup> Variant of interest Mu with E484K and N501Y mutations, and a subset with K417N, which overlaps with VOCs Beta and Gamma

**Figure 2.**
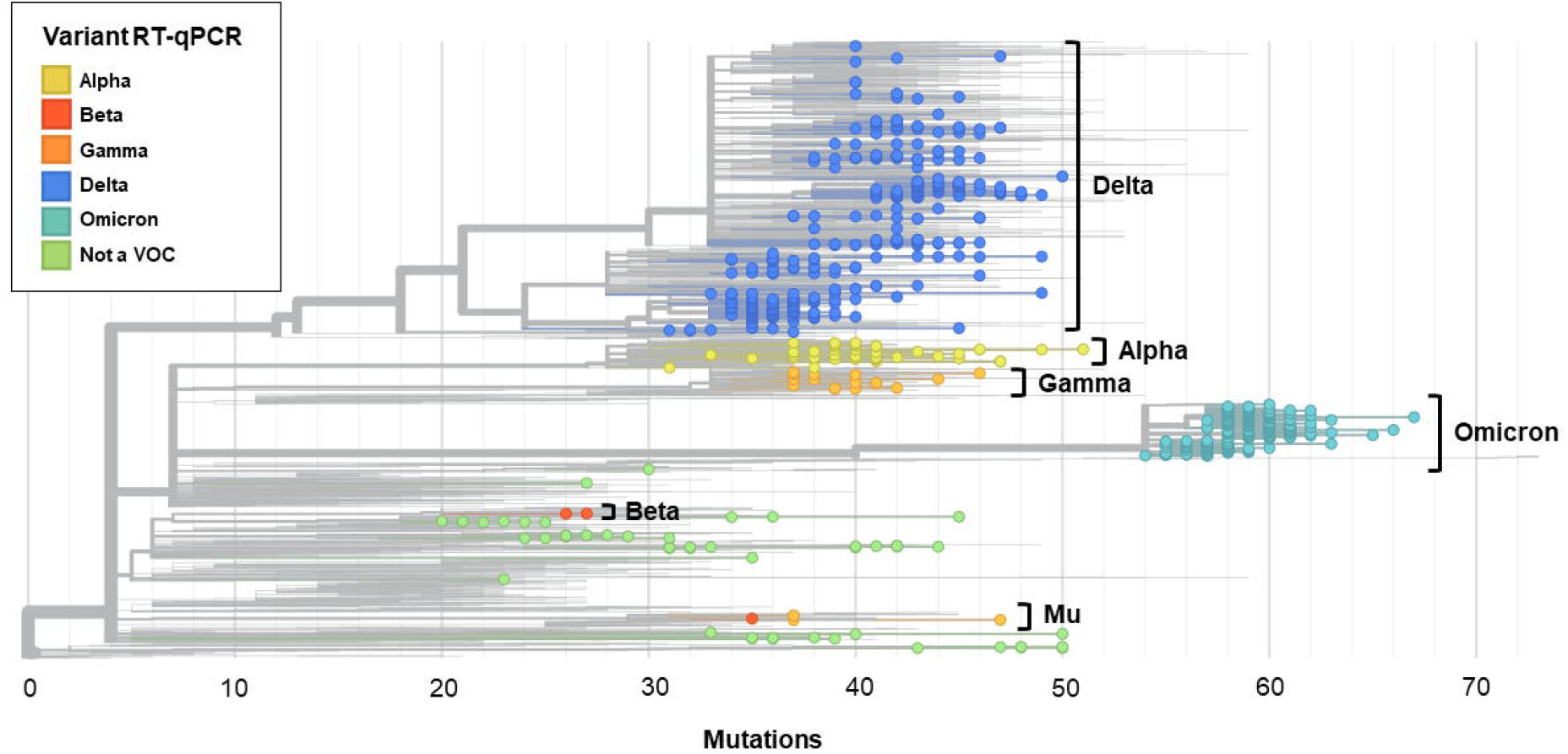
Nextclade phylogenetic tree of 3,097 SARS-CoV-2 genomes, including all 732 of the sequenced genomes from this study, and 2,365 genomes from the Nextstrain global reference tree as of February 2, 2022. The 732 included genomes are colored by RT-qPCR genotyping predicted variant type, with each circle representing a sequenced genome. Branch length corresponds to nucleotide divergence. Sequenced genomes span the breadth of the reference tree. Annotation to the right of the tree demonstrates the variant type based on whole-genome sequencing (WGS). Variant determination by RT-qPCR matched WGS except for 1 sequence typed as Beta, and 4 sequences typed as Gamma by RT-qPCR which clustered with variant of interest Mu by WGS.

## DISCUSSION

The ability to distinguish between SARS-CoV-2 VOCs is important for epidemiologic surveillance, and in certain circumstances, the care of individual COVID-19 patients. In this study, we describe a two-reaction, multiplex RT-qPCR genotyping approach that examines the spike mutations del69-70, K417N, L452R, T478K, E484K, and N501Y. This targeted mutational analysis can be used to differentiate between the WHO VOCs Alpha (B.1.1.7 and sublineages), Beta (B.1.351), Gamma (P.1 and sublineages), Delta (B.1.617.2 and sublineages), and Omicron (B.1.1.529 and sublineages), as well as identify samples which cannot be categorized into a known VOC or VOI. Because the first part of this approach, Reaction 1, has been previously described, this current study focuses on Reaction 2 and the integrated results of the two-reaction test (28). Overall, these reactions showed high concordance with WGS, demonstrating PPA 96-100% and NPA 95-100% for all targeted mutations.

Several groups have previously described similar approaches to SARS-CoV-2 variant determination by RT-qPCR and digital droplet RT-PCR, particularly for the spike del69-70, E484K, and N501Y positions (32-39). Some of these assays included additional mutation sites that were not in our study, such as spike del144 or ORF1a Δ3675–3677 (32, 38). These earlier assays, published prior to the rise of Delta, primarily targeted VOCs Alpha, Beta, and Gamma. This was then followed by a surge of reports on the detection of the Delta variant. Garson et al. utilized double-mismatch allele-specific RT-PCR at L452R and T478K to differentiate Delta variant from other VOCs in 42 UK patient samples (40). Aoki et al. described an approach that combines nested PCR along with high-resolution melting analysis at those same mutations, which was validated in a small Japanese patient cohort (41). Barua et al. used a slightly different approach, taking advantage of the difference in melting temperature of a probe targeted to Delta mutation in spike T478K compared to other variants for a Delta-specific RT-FRET-PCR assay (42). Another defining feature of VOC Delta is spike del156-157, which was the target of a Delta variant PCR test developed by Hamill et al. (43). To our knowledge, the two-reaction multiplex RT-qPCR approach outlined in this study examining six different mutation sites is the most comprehensive variant genotyping test described that can identify Alpha, Beta, Gamma, Delta, and Omicron variants.

Multiplex RT-qPCR SARS-CoV-2 genotyping takes advantage of a commonly-used molecular technique that can be implemented by laboratories using existing equipment, materials, and personnel. Because this assay is more accessible and has a more rapid turnaround time than WGS, we envision it serving a complimentary role to sequencing. The genotyping RT-qPCR can provide more detailed and up-to-date epidemiological information by increasing the sample size of categorized variants in each geographic region, and can be essential in tracking local outbreaks in areas without direct access to WGS. For individual patients, the turnaround time of several hours also allows it to directly impact clinical care. For example, VOCs show differential susceptibility to monoclonal antibody treatments, and variant reporting could include this information (Table 2) (2). Furthermore, current ongoing trials for small molecule drugs and other treatments may yield more information about variant-specific treatment strategies.

Sequencing, however, is still needed for the identification and confirmation of novel variants. This is evidenced by the 5 VOI Mu samples originally mis-identified by RT-qPCR as Gamma and Beta. The two approaches are complementary in nature, and RT-qPCR genotyping can help triage or prioritize samples for sequencing. For example, pre-screening by RT-qPCR can enrich for samples with atypical mutation patterns, lead to more efficient use of sequencing resources, and potentially, more rapid identification of new variants.

This RT-qPCR approach has several limitations as evidenced by its assay failure rate of 14% across all tested samples in our initial cohort. Because multiplex RT-qPCRs involve a mixture of multiple sets of primers and probes, they are inherently less analytically sensitive than single-target assays. For samples with RNA concentrations near the lower limits of detection, freeze-thaw cycles could impact RNA stability, and may not yield consistent results due to stochastic variation. This issue could be alleviated by implementing a Ct/RLU filter to only genotype samples most likely to yield interpretable results. Within our 1,093 sample cohort, the lower assay failure rate in samples tested in our clinical virology laboratory (4%) compared to near-care settings (35%) is likely attributable to genotyping only specimens with higher viral RNA levels. Note, however, that even with such filtering, mutation analysis by RT-qPCR remains more analytically sensitive than WGS. This study is also limited by the absence of VOC co-infections, such as with Delta and Omicron, though we anticipate that this RT-qPCR approach would be able to detect such cases. Future experiments will be required to confirm detection of VOC co-infections, including at different viral RNA levels and variant proportions.

Another limitation to this approach is the continuously evolving variant landscape which may render such a targeted assay obsolete over a relatively short period of time. First, it is important to take into account that the loss of expected internal control signal for the N501 and/or wt69-70 targets in known SARS-CoV-2 RNA positive samples is itself useful information, analogous to the widely-used Spike Gene Target Failure (36). Furthermore, the inclusion of multiple mutations in key residues that influence viral fitness and antibody escape helps guard against rapid obsolescence, as evidenced by the ability of the RT-qPCR approach to rapidly detect emergence of the Omicron variant in our population, as well as all major variant replacements that occurred in 2021. Notably, this approach also revealed emerging community transmission of BA.2 in early 2022; with K417N and wt69-70 detected in Reaction 2, and all targets including N501 not detected in Reaction 1. Nevertheless, flexibility and vigilance are required should re-design and re-validation be required as novel variants emerge.

In summary, we developed and validated a two-reaction multiplex RT-qPCR genotyping strategy that interrogates six clinically relevant mutations within the SARS-CoV-2 spike: del69-70, K417N, L452R, T478K, E484K, and N501Y. This approach allows for identification of WHO VOCs Alpha, Beta, Gamma, Delta and Omicron with excellent concordance to WGS. Overall, this method complements WGS, and is suitable for clinical decision-making, near real-time variant surveillance, and the triage of samples for sequencing.

## Data Availability

All data produced in the present study are available upon reasonable request to the authors

## ACKNOWLEDGEMENTS

We would like to thank the Stanford Clinical Virology Laboratory Staff for their excellence and perseverance in caring for our community while facing the unique challenges of this pandemic. We additionally are grateful to the Stanford Protein and Nucleic Acid Facility for their synthesis of reagents on short notice.

## Notes

### Competing Interest Statement

The authors have declared no competing interest.

### Funding Statement

This study did not receive any funding

### Author Declarations

This study was conducted with Stanford institutional review board approval (protocol 57519), and individual consent was waived.

