## Supplemental Material for "Evaluation of a Rapid and Accessible RT-qPCR Approach for SARS-CoV-2 Variant of Concern Identification"

Running Head: SARS-CoV-2 Variant of Concern Genotyping

*Authors Priscilla S.-W. Yeung and Hannah Wang contributed equally to this work. Author order was determined reverse-alphabetically.

*Assay Design*

The primers and probes for Reaction 1 have been previously reported (1). The primers and probes used for Reaction 2 are summarized in Table 1. The wt69-70 probe anneals to the same amplicon as the del69-70 probe. Primers and dual-labeled BHQ-quenched hydrolysis probes were ordered from ELIM Biopharmaceuticals (Hayward, CA, USA) or the Stanford Protein and Nucleic Acid Facility (Stanford, CA, USA), rehydrated to 100μM in Tris-EDTA buffer, and combined to create bulk primer/probe mix. We used synthetic whole-genome RNA fragments as a wild-type control (Twist Bioscience, San Francisco, CA, USA) diluted to $10^4$ copies/μL in Tris-EDTA buffer (10 mM Tris, 1 mM EDTA). For the positive control, we pooled six individual ssDNA mutant oligonucleotides (del69-70, K417N, T478K, L452R, E484K, N501Y) (Elim Biopharmaceuticals, Hayward, CA, USA) in equimolar ratios diluted to $10^4$ copies/μL each in Tris-EDTA buffer (10 mM Tris, 1 mM EDTA) (Supplemental Table 1).

For both Reaction 1 and Reaction 2, primer/probe mix (1 μL) was combined with a one-step RT-qPCR system (12.5 μL master mix + 0.5 μL *Taq* polymerase, SuperScript™ III Platinum™ One-Step qRT-PCR Kit, Invitrogen, Carlsbad), nuclease-free water (6.0 μL), and template (5.0 μL) in a 25 μL reaction. All experiments were conducted on a BioRad CFX96 real-time PCR instrument in 96-well plates (BioRad, Hercules, CA, USA). One mutant control (pooled ssDNA mutant oligonucleotides) and one wild-type control (Twist whole-genome synthetic RNA) were included in each RT-qPCR plate. Cycling conditions for Reaction 1 were: 52°C for 15:00, 94°C for 2:00, and then 45 cycles of 94°C for 00:15, 57.0°C for 00:40, and 68°C for 00:20. Annealing temperature was optimized with a temperature gradient. Cycling conditions for Reaction 2 were identical, except for the annealing temperature, which was set at 58.0°C.

Fluorescence was collected in all channels (1, T478K-FAM; 2, del60-70-HEX; 3, wt69-70-Cy3.5 (ROX); 4, K417N-CY5; 5, no probe). Fixed fluorescence thresholds of 500 relative fluorescence units ([RFU], T478K-FAM), 500 RFU (del60-70-HEX), 50 RFU (wt69-70-Cy3.5), and 300 RFU (K417N-Cy5) were used to determine the threshold cycle ($C_t$). Assay interpretation is described in detail in Table 2.

*Sample Storage and Nucleic Acid Extraction*

Samples positive for SARS-CoV-2 were stored at -80°C for up to 1 week prior to re-extraction for genotyping RT-qPCR. Total nucleic acids were extracted from 300 µL viral transport media, universal transport media, or phosphate-buffered saline, and eluted into 60 µL elution buffer on an automated platform into a 96-well plate (PerkinElmer Janus G3 Reformatter, Chemagic 360 nucleic acid extractor, and Chemagic Viral DNA/RNA 300 Kit). Eluate plates were stored for up to 6 months at 4°C between setup of genotyping RT-qPCR Reaction 1 and Reaction 2.

*Clinical Specimen NAAT Platforms*

Prior to genotyping RT-qPCR, initial respiratory SARS-CoV-2 NAAT was conducted on a variety of platforms (Table 1) (2-4). These included: 1) a previously-described laboratory-developed reverse transcription quantitative polymerase chain reaction (RT-qPCR) targeting the envelope gene (*E* gene) on the Rotor-Gene Q (Qiagen, Germantown, MD) (2-4); 2) PerkinElmer RT-qPCR assay targeting open reading frame 1ab (ORF1ab) and nucleocapsid gene ([*N* gene] PerkinElmer, San Jose, CA); 3) Panther Fusion SARS-CoV-2 (Hologic, Marlborough, MA) RT-qPCR targeting ORF1ab; 4) Aptima SARS-CoV2 (Panther System, Hologic) transcription mediated amplification targeting ORF1ab; 5) GeneXpert Xpress SARS-CoV-2 (Cepheid, Sunnyvale, CA) RT-qPCR targeting the *E* and *N* genes; 6) cobas Liat SARS-CoV-2 & Influenza A/B (Roche, Indianapolis, IN), RT-qPCR targeting ORF1ab and *N* gene; 7) e-Plex SARS-CoV-2 (Genmark, Carlsbad, CA), RT-PCR targeting the *N* gene. All specimens testing positive for SARS-CoV-2 by NAAT with RT-qPCR $C_t \leq 30$ or transcription-mediated amplification relative light units (RLU) ≥ 1,100 during this period were subject to multiplex allele-specific genotyping RT-qPCR. A subset of included specimens tested by rapid NAATs (Cepheid GeneXpert, Roche Liat, Genmark Eplex) did not have $C_t$ values available and were included irrespective of viral load. All assays were conducted according to manufacturer and emergency authorization instructions.

*Analytical Performance*

To determine the lower limit of detection (LLOD), the pool of six mutant ssDNA oligonucleotides described above was diluted to 100 copies/µL template, 10 copies/µL, 5 copies/µL, and 1 copies/µL in Tris-EDTA buffer in replicates of 20. Any amplification crossing the fluorescence threshold was regarded as detection. The 95% LLOD was determined by fitting these data to a probit regression curve. The respective 95% LLODs for the del69-70, K417N, and T478K targets were 14.8 (95% CI 10.5-27.2), 16.4 (11.3-44.3), and 2.1 (2.1-8.1) copies/μL template respectively (Supplemental Table 2). We observed no non-specific wt69-70 (Cy3.5) amplification even at high ($10^6$ copies/μL) mutant ssDNA copy number; similarly, we observed no non-specific del69-70, K417N, and T478K non-specific amplification at high concentration ($10^6$ copies/μL) of the wild-type TWIST synthetic RNA. Precision was not assessed for this qualitative assay.

**Supplemental Figure 1.** In silico analysis primer/probes, per GISAID Nextclade sequences downloaded December 5, 2021. Green shading indicates >95% of sequences from that variant designation have perfect homology to a primer/probe sequence expected to be detected in this variant. Red shading indicates <95% of sequences are homologous to a probe sequence expected to be detected in this variant, and probe dropout or diminished efficiency of binding is expected to occur. Yellow shading indicates >5% of sequences have unexpected probe homology to the mutation of interest.

| Primer/Probe | Alpha | Beta | Delta | Gamma | Omicron | Lambda | Mu | Non-VOC/VOI |
|---|---|---|---|---|---|---|---|---|
| **L452R_FWD** | 100% (n=164/164) | 100% (n=35/35) | 99.7% (n=2082/2089) | 98.8% (n=81/82) | 100% (n=14/14) | 100% (n=22/22) | 100% (n=22/22) | 99.7% (n=950/953) |
| **L452R_REV** | 99.4% (n=163/164) | 100% (n=35/35) | 99.1% (n=2069/2087) | 100% (n=82/82) | 0% (n=0/14) | 100% (n=22/22) | 100% (n=22/22) | 99.6% (n=941/945) |
| **L452R_mt_HEX** | 0% (n=0/163) | 0% (n=0/35) | 99.3% (n=2073/2088) | 0% (n=0/81) | 0% (n=0/14) | 0% (n=0/22) | 0% (n=0/22) | 9.2% (n=87/945) |
| **E484K_FWD_V2** | 98.2% (n=160/163) | 100% (n=35/35) | 99.7% (n=2081/2088) | 97.5% (n=79/81) | 100% (n=17/17) | 100% (n=22/22) | 100% (n=22/22) | 99.3% (n=943/950) |
| **E484K_REV_V2** | 100% (n=163/163) | 100% (n=35/35) | 99.7% (n=2024/2030) | 98.8% (n=81/82) | 100% (n=19/19) | 100% (n=24/24) | 95.5% (n=21/22) | 99.9% (n=943/944) |
| **E484K_mt_CY5** | 1.8% (n=3/165) | 97.2% (n=35/36) | 0.1% (n=2/2089) | 98.8% (n=79/80) | 0% (n=0/18) | 0% (n=0/24) | 100% (n=21/21) | 6.9% (n=65/948) |
| **N501Y_mt_FAM** | 100% (n=161/161) | 94.4% (n=34/36) | 0% (n=0/2043) | 100% (n=81/81) | 0% (n=0/18) | 0% (n=0/24) | 100% (n=21/21) | 1.4% (n=13/946) |
| **N501Y_WT_CY3.5** | 0% (n=0/161) | 5.6% (n=2/36) | 99.7% (n=2037/2043) | 0% (n=0/81) | 0% (n=0/18) | 100% (n=24/24) | 0% (n=0/21) | 97.4% (n=921/946) |
| **del69.70_FWD_V2** | 100% (n=169/169) | 97.4% (n=38/39) | 99.5% (n=1714/1722) | 100% (n=81/81) | 100% (n=19/19) | 100% (n=24/24) | 100% (n=22/22) | 99.8% (n=968/970) |
| **del69.70_REV_V2** | 99.4% (n=170/171) | 100% (n=39/39) | 99.3% (n=1862/1876) | 100% (n=81/81) | 100% (n=19/19) | 100% (n=24/24) | 100% (n=22/22) | 96.8% (n=945/976) |
| **del69.70_mt_HEX** | 99.4% (n=166/167) | 0% (n=0/39) | 0.2% (n=4/1747) | 1.2% (n=1/81) | 0% (n=0/17) | 0% (n=0/24) | 0% (n=0/22) | 1.8% (n=17/970) |
| **wt69.70_wt_Cy3.5** | 0.6% (n=1/167) | 100% (n=39/39) | 97% (n=1704/1757) | 98.8% (n=80/81) | 0% (n=0/17) | 95.8% (n=23/24) | 100% (n=22/22) | 95.2% (n=923/970) |
| **K417N_FWD_V2** | 99.4% (n=169/170) | 100% (n=39/39) | 99.6% (n=2109/2118) | 97.6% (n=81/83) | 100% (n=14/14) | 100% (n=22/22) | 100% (n=22/22) | 99.6% (n=989/993) |
| **K417N_REV_V2** | 100% (n=172/172) | 100% (n=38/38) | 99.5% (n=2109/2120) | 100% (n=83/83) | 100% (n=18/18) | 95.8% (n=23/24) | 100% (n=22/22) | 99.9% (n=980/981) |
| **K417N_mt_CY5_V4** | 0% (n=0/170) | 97.4% (n=38/39) | 0% (n=1/2120) | 0% (n=0/82) | 100% (n=14/14) | 0% (n=0/22) | 0% (n=0/22) | 0.2% (n=2/986) |
| **T478K_FWD** | 97.6% (n=162/166) | 100% (n=36/36) | 99.6% (n=2081/2090) | 97.5% (n=79/81) | 100% (n=18/18) | 0% (n=0/24) | 100% (n=22/22) | 99.5% (n=944/949) |
| **T478K_REV** | 100% (n=164/164) | 100% (n=35/35) | 99.7% (n=2081/2088) | 98.8% (n=81/82) | 100% (n=14/14) | 100% (n=22/22) | 100% (n=22/22) | 99.7% (n=951/954) |
| **T478K_mt_FAM** | 0% (n=0/164) | 0% (n=0/36) | 97.9% (n=2049/2092) | 0% (n=0/82) | 0% (n=0/18) | 0% (n=0/24) | 0% (n=0/22) | 7.7% (n=73/949) |

**Supplemental Figure 2.** Flow chart of specimens sent for nucleic acid amplification tests (NAAT) during the initial study period and number of specimens ultimately genotyped by the two-reaction multiplex RT-qPCR and sequenced.

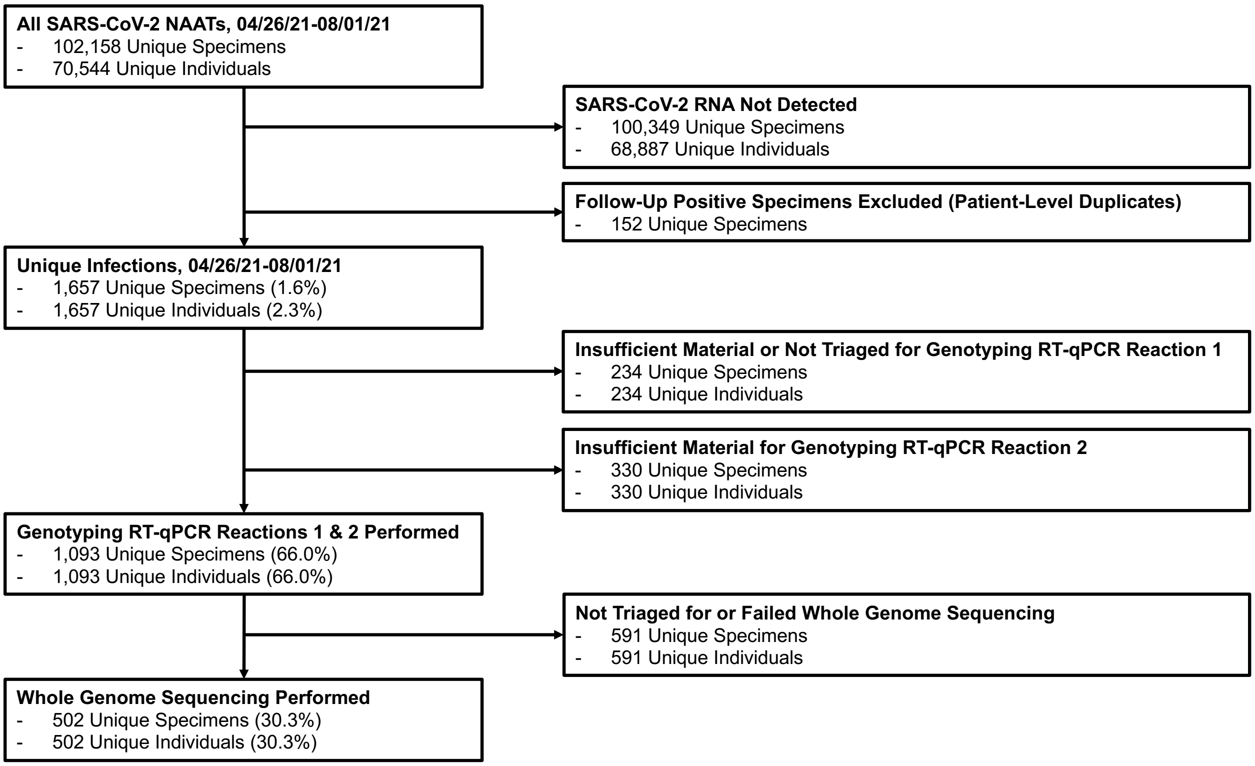

**Supplemental Table 1.** Sequences of Mutant Oligonucleotide Controls

| Name[a] | Position and base change[b] | Length (bp) | Sequence (5' → 3') |
|---|---|---|---|
| ssDNA_L452R_MT | 22917 T>G | 78 | TCTCTCAAAAGGTTTGAGATTAGACTTCCTAAACAATCTATACCGGTAATT ATAATTACCACCAACCTTAGAATCAAG |
| ssDNA_del69-70_MT | del21765-70 | 113 | ACATTCAACTCAGGACTTGTTCTTACCTTTCTTTTCCAATGTTACTTGGTT CCATGCTATCTCTGGGACCAATGGTACTAAGAGGTTTGATAACCCTGTCC TACCATTTAATG |
| ssDNA_K417N_MT | 22813G>T | 101 | CATTTGTAATTAGAGGTGATGAAGTCAGACAAATCGCTCCAGGGCAAACT GGAAATATTGCTGATTATAATTATAAATTACCAGATGATTTTACAGGCTGC |
| ssDNA_T478K_MT | 22995 C>A | 117 | ATTGTAAAGGAAAGTAACAATTAAAACCTTCAACACCATTACAAGGTTTGC TACCGGCCTGATAGATTTCAGTTGAAATATCTCTCTCAAAAGGTTTGAGA TTAGACTTCCTAAACA |
| ssDNA_E484K_MT | 23012G>A | 133 | CTGAAATCTATCAGGCCGGTAGCACACCTTGTAATGGTGTTAAAGGTTTT AATTGTTACTTTCCTTTACAATCATATGGTTTCCAACCCACTTATGGTGTT GGTTACCAACCATACAGAGTAGTAGTACTTTC |
| ssDNA_N501Y_MT | 23063A>T | 134 | GTTTTAATTGTTACTTTCCTTTACAATCATATGGTTTCCAACCCACTTATG GTGTTGGTTACCAACCATACAGAGTAGTAGTACTTTCTTTTGAACTTCTAC ATGCACCAGCAACTGTTTGTGGACCTAAAAAG |

MT, mutant; bp, base pairs

[a] Individual ssDNA oligonucleotides were pooled in equimolar ratios diluted to $10^4$ copies/µL each in Tris-EDTA buffer to create the mutant control.

[b] Position and sequence based on Genbank NC_045512

**Supplemental Table 2.** Lower Limit of Detection for Reaction 2 mutation targets

| **Concentration (copies/μL template)** | **Number of Detected Replicates** | | |
|---|---|---|---|
| | **del69-70-mt (HEX)** | **K417N-mt (CY5)** | **T478K-mt (FAM)** |
| 1.0 | 5/20 | 6/20 | 10/20 |
| 5.0 | 12/20 | 13/20 | 20/20 |
| 10.0 | 16/20 | 15/20 | 20/20 |
| 100.0 | 20/20 | 20/20 | 20/20 |
| **95% LLOD (95% CI)** | **14.8 (10.5-27.2)** | **16.4 (11.3-44.3)** | **2.1 (2.1-8.1)** |

**Supplemental Table 3.** GISAID Accession Numbers for Sequenced Study Samples (n=547)

| Predicted Variant Type by Genotyping RT-qPCR | Variant Type by Whole Genome Sequencing | PANGO Lineage[a] | GISAID Accession |
|---|---|---|---|
| Alpha | Alpha | B.1.1.7 | EPI_ISL_3050185 |
| Alpha | Alpha | B.1.1.7 | EPI_ISL_3050188 |
| Alpha | Alpha | B.1.1.7 | EPI_ISL_3050190 |
| Alpha | Alpha | B.1.1.7 | EPI_ISL_3050194 |
| Alpha | Alpha | B.1.1.7 | EPI_ISL_3050198 |
| Alpha | Alpha | B.1.1.7 | EPI_ISL_3050205 |
| Alpha | Alpha | B.1.1.7 | EPI_ISL_3050207 |
| Alpha | Alpha | B.1.1.7 | EPI_ISL_4072053 |
| Alpha | Alpha | B.1.1.7 | EPI_ISL_4072055 |
| Alpha | Alpha | B.1.1.7 | EPI_ISL_4072056 |
| Alpha | Alpha | B.1.1.7 | EPI_ISL_4072057 |
| Alpha | Alpha | B.1.1.7 | EPI_ISL_4072058 |
| Alpha | Alpha | B.1.1.7 | EPI_ISL_4072059 |
| Alpha | Alpha | B.1.1.7 | EPI_ISL_4072060 |
| Alpha | Alpha | B.1.1.7 | EPI_ISL_4072061 |
| Alpha | Alpha | B.1.1.7 | EPI_ISL_4072062 |
| Alpha | Alpha | B.1.1.7 | EPI_ISL_4072063 |
| Alpha | Alpha | B.1.1.7 | EPI_ISL_4072064 |
| Alpha | Alpha | B.1.1.7 | EPI_ISL_4072065 |
| Alpha | Alpha | B.1.1.7 | EPI_ISL_4072068 |
| Alpha | Alpha | B.1.1.7 | EPI_ISL_4072069 |
| Alpha | Alpha | B.1.1.7 | EPI_ISL_4496848 |
| Alpha | Alpha | B.1.1.7 | EPI_ISL_4496849 |
| Alpha | Alpha | B.1.1.7 | EPI_ISL_4496850 |
| Alpha | Alpha | B.1.1.7 | EPI_ISL_4496851 |
| Alpha | Alpha | B.1.1.7 | EPI_ISL_4496857 |
| Alpha | Alpha | B.1.1.7 | EPI_ISL_4496858 |
| Alpha | Alpha | B.1.1.7 | EPI_ISL_4496860 |
| Alpha | Alpha | B.1.1.7 | EPI_ISL_4496861 |
| Alpha | Alpha | B.1.1.7 | EPI_ISL_4496865 |
| Alpha | Alpha | B.1.1.7 | EPI_ISL_4496867 |
| Alpha | Alpha | B.1.1.7 | EPI_ISL_4496868 |
| Alpha | Alpha | B.1.1.7 | EPI_ISL_4496905 |
| Alpha | Alpha | B.1.1.7 | EPI_ISL_4496906 |
| Alpha | Alpha | B.1.1.7 | EPI_ISL_4496907 |
| Alpha | Alpha | B.1.1.7 | EPI_ISL_4496908 |
| Alpha | Alpha | B.1.1.7 | EPI_ISL_4496909 |
| Alpha | Alpha | Q.3 | EPI_ISL_3050181 |
| Alpha | Alpha | Q.3 | EPI_ISL_3050189 |
| Alpha | Alpha | Q.3 | EPI_ISL_4072066 |
| Alpha | Alpha | Q.3 | EPI_ISL_4072067 |
| Alpha | Alpha | Q.3 | EPI_ISL_4072071 |
| Alpha | Alpha | Q.3 | EPI_ISL_4072072 |
| Beta | Beta | B.1.351 | EPI_ISL_4496869 |
| Beta | Beta | B.1.351 | EPI_ISL_4496898 |
| Beta | Not a VOC | B.1.621 | EPI_ISL_3236162 |
| Delta | Delta | AY.1 | EPI_ISL_2987140 |
| Delta | Delta | AY.1 | EPI_ISL_2987141 |
| Delta | Delta | AY.1 | EPI_ISL_2987142 |
| Delta | Delta | AY.1 | EPI_ISL_3050224 |
| Delta | Delta | AY.1 | EPI_ISL_3236110 |
| Delta | Delta | AY.1 | EPI_ISL_3236112 |

| | | | |
|---|---|---|---|
| Delta | Delta | AY.1 | EPI_ISL_3236124 |
| Delta | Delta | AY.1 | EPI_ISL_3236131 |
| Delta | Delta | AY.1 | EPI_ISL_3236146 |
| Delta | Delta | AY.1 | EPI_ISL_3236161 |
| Delta | Delta | AY.1 | EPI_ISL_3236167 |
| Delta | Delta | AY.1 | EPI_ISL_3390756 |
| Delta | Delta | AY.1 | EPI_ISL_3916305 |
| Delta | Delta | AY.1 | EPI_ISL_3916308 |
| Delta | Delta | AY.1 | EPI_ISL_3916328 |
| Delta | Delta | AY.1 | EPI_ISL_3916330 |
| Delta | Delta | AY.1 | EPI_ISL_3916341 |
| Delta | Delta | AY.1 | EPI_ISL_3916368 |
| Delta | Delta | AY.1 | EPI_ISL_3916373 |
| Delta | Delta | AY.1 | EPI_ISL_3916390 |
| Delta | Delta | AY.100 | EPI_ISL_3390725 |
| Delta | Delta | AY.100 | EPI_ISL_3538733 |
| Delta | Delta | AY.100 | EPI_ISL_3916354 |
| Delta | Delta | AY.103 | EPI_ISL_3050256 |
| Delta | Delta | AY.103 | EPI_ISL_3236104 |
| Delta | Delta | AY.103 | EPI_ISL_3236145 |
| Delta | Delta | AY.103 | EPI_ISL_3390747 |
| Delta | Delta | AY.103 | EPI_ISL_3538732 |
| Delta | Delta | AY.103 | EPI_ISL_3538737 |
| Delta | Delta | AY.103 | EPI_ISL_3538762 |
| Delta | Delta | AY.103 | EPI_ISL_3538764 |
| Delta | Delta | AY.103 | EPI_ISL_3538768 |
| Delta | Delta | AY.103 | EPI_ISL_3722174 |
| Delta | Delta | AY.103 | EPI_ISL_3722189 |
| Delta | Delta | AY.103 | EPI_ISL_3722193 |
| Delta | Delta | AY.103 | EPI_ISL_3722208 |
| Delta | Delta | AY.103 | EPI_ISL_3916283 |
| Delta | Delta | AY.103 | EPI_ISL_3916287 |
| Delta | Delta | AY.103 | EPI_ISL_3916294 |
| Delta | Delta | AY.103 | EPI_ISL_3916312 |
| Delta | Delta | AY.103 | EPI_ISL_3916320 |
| Delta | Delta | AY.103 | EPI_ISL_3916339 |
| Delta | Delta | AY.103 | EPI_ISL_3916345 |
| Delta | Delta | AY.103 | EPI_ISL_3916349 |
| Delta | Delta | AY.103 | EPI_ISL_3916351 |
| Delta | Delta | AY.103 | EPI_ISL_3916362 |
| Delta | Delta | AY.103 | EPI_ISL_3916366 |
| Delta | Delta | AY.103 | EPI_ISL_3916367 |
| Delta | Delta | AY.103 | EPI_ISL_3916391 |
| Delta | Delta | AY.110 | EPI_ISL_3050183 |
| Delta | Delta | AY.110 | EPI_ISL_3236144 |
| Delta | Delta | AY.110 | EPI_ISL_3236164 |
| Delta | Delta | AY.110 | EPI_ISL_3236165 |
| Delta | Delta | AY.110 | EPI_ISL_3236171 |
| Delta | Delta | AY.110 | EPI_ISL_3390748 |
| Delta | Delta | AY.110 | EPI_ISL_3538729 |
| Delta | Delta | AY.110 | EPI_ISL_4072017 |
| Delta | Delta | AY.110 | EPI_ISL_4496828 |
| Delta | Delta | AY.114 | EPI_ISL_3722179 |
| Delta | Delta | AY.116.1 | EPI_ISL_3236127 |
| Delta | Delta | AY.116.1 | EPI_ISL_3538748 |
| Delta | Delta | AY.118 | EPI_ISL_3050199 |

| | | | |
|---|---|---|---|
| Delta | Delta | AY.118 | EPI_ISL_3050208 |
| Delta | Delta | AY.118 | EPI_ISL_3236107 |
| Delta | Delta | AY.118 | EPI_ISL_3236169 |
| Delta | Delta | AY.118 | EPI_ISL_3916369 |
| Delta | Delta | AY.119 | EPI_ISL_3236172 |
| Delta | Delta | AY.119 | EPI_ISL_3722212 |
| Delta | Delta | AY.119 | EPI_ISL_3916299 |
| Delta | Delta | AY.119 | EPI_ISL_4072028 |
| Delta | Delta | AY.120.1 | EPI_ISL_3916300 |
| Delta | Delta | AY.121 | EPI_ISL_3236111 |
| Delta | Delta | AY.121 | EPI_ISL_3236115 |
| Delta | Delta | AY.121 | EPI_ISL_3538735 |
| Delta | Delta | AY.122 | EPI_ISL_3236149 |
| Delta | Delta | AY.122 | EPI_ISL_3236181 |
| Delta | Delta | AY.122 | EPI_ISL_3916311 |
| Delta | Delta | AY.122 | EPI_ISL_4496829 |
| Delta | Delta | AY.122 | EPI_ISL_4496830 |
| Delta | Delta | AY.126 | EPI_ISL_3050182 |
| Delta | Delta | AY.126 | EPI_ISL_3236175 |
| Delta | Delta | AY.13 | EPI_ISL_3050253 |
| Delta | Delta | AY.13 | EPI_ISL_3236106 |
| Delta | Delta | AY.13 | EPI_ISL_3236109 |
| Delta | Delta | AY.13 | EPI_ISL_3236113 |
| Delta | Delta | AY.13 | EPI_ISL_3236114 |
| Delta | Delta | AY.13 | EPI_ISL_3236130 |
| Delta | Delta | AY.13 | EPI_ISL_3236141 |
| Delta | Delta | AY.13 | EPI_ISL_3236156 |
| Delta | Delta | AY.13 | EPI_ISL_3236158 |
| Delta | Delta | AY.13 | EPI_ISL_3236160 |
| Delta | Delta | AY.13 | EPI_ISL_3236179 |
| Delta | Delta | AY.13 | EPI_ISL_3236180 |
| Delta | Delta | AY.13 | EPI_ISL_3236183 |
| Delta | Delta | AY.13 | EPI_ISL_3236185 |
| Delta | Delta | AY.13 | EPI_ISL_3390740 |
| Delta | Delta | AY.13 | EPI_ISL_3390743 |
| Delta | Delta | AY.13 | EPI_ISL_3390744 |
| Delta | Delta | AY.13 | EPI_ISL_3538753 |
| Delta | Delta | AY.13 | EPI_ISL_3538754 |
| Delta | Delta | AY.13 | EPI_ISL_3538756 |
| Delta | Delta | AY.13 | EPI_ISL_3722199 |
| Delta | Delta | AY.13 | EPI_ISL_3916270 |
| Delta | Delta | AY.13 | EPI_ISL_3916319 |
| Delta | Delta | AY.13 | EPI_ISL_3916353 |
| Delta | Delta | AY.13 | EPI_ISL_3916386 |
| Delta | Delta | AY.13 | EPI_ISL_4072014 |
| Delta | Delta | AY.13 | EPI_ISL_4072016 |
| Delta | Delta | AY.13 | EPI_ISL_4072024 |
| Delta | Delta | AY.13 | EPI_ISL_4072025 |
| Delta | Delta | AY.13 | EPI_ISL_4072026 |
| Delta | Delta | AY.13 | EPI_ISL_4496853 |
| Delta | Delta | AY.13 | EPI_ISL_4496885 |
| Delta | Delta | AY.14 | EPI_ISL_3050184 |
| Delta | Delta | AY.14 | EPI_ISL_3050195 |
| Delta | Delta | AY.14 | EPI_ISL_3050196 |
| Delta | Delta | AY.14 | EPI_ISL_3050201 |
| Delta | Delta | AY.14 | EPI_ISL_3050202 |

| | | | |
|---|---|---|---|
| Delta | Delta | AY.14 | EPI_ISL_3050211 |
| Delta | Delta | AY.14 | EPI_ISL_3236105 |
| Delta | Delta | AY.14 | EPI_ISL_3236108 |
| Delta | Delta | AY.14 | EPI_ISL_3236117 |
| Delta | Delta | AY.14 | EPI_ISL_3236119 |
| Delta | Delta | AY.14 | EPI_ISL_3236121 |
| Delta | Delta | AY.14 | EPI_ISL_3236123 |
| Delta | Delta | AY.14 | EPI_ISL_3236125 |
| Delta | Delta | AY.14 | EPI_ISL_3236134 |
| Delta | Delta | AY.14 | EPI_ISL_3236135 |
| Delta | Delta | AY.14 | EPI_ISL_3236138 |
| Delta | Delta | AY.14 | EPI_ISL_3236148 |
| Delta | Delta | AY.14 | EPI_ISL_3236174 |
| Delta | Delta | AY.14 | EPI_ISL_3236177 |
| Delta | Delta | AY.14 | EPI_ISL_3390726 |
| Delta | Delta | AY.14 | EPI_ISL_3390732 |
| Delta | Delta | AY.14 | EPI_ISL_3390734 |
| Delta | Delta | AY.14 | EPI_ISL_3390737 |
| Delta | Delta | AY.14 | EPI_ISL_3390753 |
| Delta | Delta | AY.14 | EPI_ISL_3538738 |
| Delta | Delta | AY.14 | EPI_ISL_3538739 |
| Delta | Delta | AY.14 | EPI_ISL_3538741 |
| Delta | Delta | AY.14 | EPI_ISL_3538744 |
| Delta | Delta | AY.14 | EPI_ISL_3538750 |
| Delta | Delta | AY.14 | EPI_ISL_3538751 |
| Delta | Delta | AY.14 | EPI_ISL_3538757 |
| Delta | Delta | AY.14 | EPI_ISL_3538760 |
| Delta | Delta | AY.14 | EPI_ISL_3538766 |
| Delta | Delta | AY.14 | EPI_ISL_3722184 |
| Delta | Delta | AY.14 | EPI_ISL_3722185 |
| Delta | Delta | AY.14 | EPI_ISL_3722186 |
| Delta | Delta | AY.14 | EPI_ISL_3722187 |
| Delta | Delta | AY.14 | EPI_ISL_3722194 |
| Delta | Delta | AY.14 | EPI_ISL_3722196 |
| Delta | Delta | AY.14 | EPI_ISL_3722197 |
| Delta | Delta | AY.14 | EPI_ISL_3722200 |
| Delta | Delta | AY.14 | EPI_ISL_3916275 |
| Delta | Delta | AY.14 | EPI_ISL_3916296 |
| Delta | Delta | AY.14 | EPI_ISL_3916298 |
| Delta | Delta | AY.14 | EPI_ISL_3916309 |
| Delta | Delta | AY.14 | EPI_ISL_3916322 |
| Delta | Delta | AY.14 | EPI_ISL_3916323 |
| Delta | Delta | AY.14 | EPI_ISL_3916327 |
| Delta | Delta | AY.14 | EPI_ISL_3916336 |
| Delta | Delta | AY.14 | EPI_ISL_3916350 |
| Delta | Delta | AY.14 | EPI_ISL_3916357 |
| Delta | Delta | AY.14 | EPI_ISL_3916365 |
| Delta | Delta | AY.14 | EPI_ISL_3916370 |
| Delta | Delta | AY.14 | EPI_ISL_3916371 |
| Delta | Delta | AY.14 | EPI_ISL_3916372 |
| Delta | Delta | AY.14 | EPI_ISL_3916377 |
| Delta | Delta | AY.14 | EPI_ISL_3916383 |
| Delta | Delta | AY.14 | EPI_ISL_3916385 |
| Delta | Delta | AY.14 | EPI_ISL_4072027 |
| Delta | Delta | AY.19 | EPI_ISL_3916364 |
| Delta | Delta | AY.2 | EPI_ISL_3050234 |

| | | | |
|---|---|---|---|
| Delta | Delta | AY.2 | EPI_ISL_3050247 |
| Delta | Delta | AY.2 | EPI_ISL_3236142 |
| Delta | Delta | AY.2 | EPI_ISL_3236159 |
| Delta | Delta | AY.2 | EPI_ISL_3390749 |
| Delta | Delta | AY.20 | EPI_ISL_3236129 |
| Delta | Delta | AY.20 | EPI_ISL_3538740 |
| Delta | Delta | AY.20 | EPI_ISL_3916290 |
| Delta | Delta | AY.20 | EPI_ISL_3916335 |
| Delta | Delta | AY.20 | EPI_ISL_3916375 |
| Delta | Delta | AY.23 | EPI_ISL_3390755 |
| Delta | Delta | AY.25 | EPI_ISL_3050252 |
| Delta | Delta | AY.25 | EPI_ISL_3236184 |
| Delta | Delta | AY.25 | EPI_ISL_3390758 |
| Delta | Delta | AY.25 | EPI_ISL_3538736 |
| Delta | Delta | AY.25 | EPI_ISL_3722205 |
| Delta | Delta | AY.25 | EPI_ISL_3916276 |
| Delta | Delta | AY.25 | EPI_ISL_3916352 |
| Delta | Delta | AY.25.1 | EPI_ISL_3236133 |
| Delta | Delta | AY.25.1 | EPI_ISL_3236137 |
| Delta | Delta | AY.25.1 | EPI_ISL_3390730 |
| Delta | Delta | AY.25.1 | EPI_ISL_3390745 |
| Delta | Delta | AY.25.1 | EPI_ISL_3390746 |
| Delta | Delta | AY.25.1 | EPI_ISL_3390750 |
| Delta | Delta | AY.25.1 | EPI_ISL_3390751 |
| Delta | Delta | AY.25.1 | EPI_ISL_3390752 |
| Delta | Delta | AY.25.1 | EPI_ISL_3538730 |
| Delta | Delta | AY.25.1 | EPI_ISL_3538765 |
| Delta | Delta | AY.25.1 | EPI_ISL_3722206 |
| Delta | Delta | AY.25.1 | EPI_ISL_3916269 |
| Delta | Delta | AY.25.1 | EPI_ISL_3916272 |
| Delta | Delta | AY.25.1 | EPI_ISL_3916278 |
| Delta | Delta | AY.25.1 | EPI_ISL_3916304 |
| Delta | Delta | AY.25.1 | EPI_ISL_3916310 |
| Delta | Delta | AY.25.1 | EPI_ISL_3916314 |
| Delta | Delta | AY.25.1 | EPI_ISL_3916324 |
| Delta | Delta | AY.25.1 | EPI_ISL_3916325 |
| Delta | Delta | AY.25.1 | EPI_ISL_3916333 |
| Delta | Delta | AY.25.1 | EPI_ISL_3916334 |
| Delta | Delta | AY.25.1 | EPI_ISL_3916338 |
| Delta | Delta | AY.25.1 | EPI_ISL_3916347 |
| Delta | Delta | AY.25.1 | EPI_ISL_3916358 |
| Delta | Delta | AY.25.1 | EPI_ISL_3916389 |
| Delta | Delta | AY.26 | EPI_ISL_3050193 |
| Delta | Delta | AY.26 | EPI_ISL_3050246 |
| Delta | Delta | AY.26 | EPI_ISL_3236136 |
| Delta | Delta | AY.26 | EPI_ISL_3236178 |
| Delta | Delta | AY.26 | EPI_ISL_3390731 |
| Delta | Delta | AY.26 | EPI_ISL_3390735 |
| Delta | Delta | AY.26 | EPI_ISL_3390754 |
| Delta | Delta | AY.26 | EPI_ISL_3722182 |
| Delta | Delta | AY.26 | EPI_ISL_3722192 |
| Delta | Delta | AY.26 | EPI_ISL_3722201 |
| Delta | Delta | AY.26 | EPI_ISL_3722209 |
| Delta | Delta | AY.26 | EPI_ISL_3722210 |
| Delta | Delta | AY.26 | EPI_ISL_3916297 |
| Delta | Delta | AY.26 | EPI_ISL_3916301 |

| | | | |
|---|---|---|---|
| Delta | Delta | AY.26 | EPI_ISL_4072040 |
| Delta | Delta | AY.3 | EPI_ISL_3538763 |
| Delta | Delta | AY.3 | EPI_ISL_3722190 |
| Delta | Delta | AY.3 | EPI_ISL_3916280 |
| Delta | Delta | AY.3 | EPI_ISL_3916302 |
| Delta | Delta | AY.3 | EPI_ISL_3916376 |
| Delta | Delta | AY.35 | EPI_ISL_3236173 |
| Delta | Delta | AY.35 | EPI_ISL_3236176 |
| Delta | Delta | AY.4 | EPI_ISL_3916286 |
| Delta | Delta | AY.43 | EPI_ISL_3916342 |
| Delta | Delta | AY.43 | EPI_ISL_3916388 |
| Delta | Delta | AY.44 | EPI_ISL_3050200 |
| Delta | Delta | AY.44 | EPI_ISL_3236102 |
| Delta | Delta | AY.44 | EPI_ISL_3236103 |
| Delta | Delta | AY.44 | EPI_ISL_3236116 |
| Delta | Delta | AY.44 | EPI_ISL_3236118 |
| Delta | Delta | AY.44 | EPI_ISL_3236120 |
| Delta | Delta | AY.44 | EPI_ISL_3236128 |
| Delta | Delta | AY.44 | EPI_ISL_3236153 |
| Delta | Delta | AY.44 | EPI_ISL_3236154 |
| Delta | Delta | AY.44 | EPI_ISL_3236155 |
| Delta | Delta | AY.44 | EPI_ISL_3236166 |
| Delta | Delta | AY.44 | EPI_ISL_3390728 |
| Delta | Delta | AY.44 | EPI_ISL_3390729 |
| Delta | Delta | AY.44 | EPI_ISL_3390736 |
| Delta | Delta | AY.44 | EPI_ISL_3390739 |
| Delta | Delta | AY.44 | EPI_ISL_3390741 |
| Delta | Delta | AY.44 | EPI_ISL_3390742 |
| Delta | Delta | AY.44 | EPI_ISL_3538728 |
| Delta | Delta | AY.44 | EPI_ISL_3538731 |
| Delta | Delta | AY.44 | EPI_ISL_3538734 |
| Delta | Delta | AY.44 | EPI_ISL_3538743 |
| Delta | Delta | AY.44 | EPI_ISL_3538745 |
| Delta | Delta | AY.44 | EPI_ISL_3538747 |
| Delta | Delta | AY.44 | EPI_ISL_3538749 |
| Delta | Delta | AY.44 | EPI_ISL_3538755 |
| Delta | Delta | AY.44 | EPI_ISL_3538758 |
| Delta | Delta | AY.44 | EPI_ISL_3538759 |
| Delta | Delta | AY.44 | EPI_ISL_3538761 |
| Delta | Delta | AY.44 | EPI_ISL_3538767 |
| Delta | Delta | AY.44 | EPI_ISL_3722175 |
| Delta | Delta | AY.44 | EPI_ISL_3722177 |
| Delta | Delta | AY.44 | EPI_ISL_3722180 |
| Delta | Delta | AY.44 | EPI_ISL_3722181 |
| Delta | Delta | AY.44 | EPI_ISL_3722183 |
| Delta | Delta | AY.44 | EPI_ISL_3722188 |
| Delta | Delta | AY.44 | EPI_ISL_3722191 |
| Delta | Delta | AY.44 | EPI_ISL_3722195 |
| Delta | Delta | AY.44 | EPI_ISL_3722202 |
| Delta | Delta | AY.44 | EPI_ISL_3722203 |
| Delta | Delta | AY.44 | EPI_ISL_3722204 |
| Delta | Delta | AY.44 | EPI_ISL_3722211 |
| Delta | Delta | AY.44 | EPI_ISL_3722214 |
| Delta | Delta | AY.44 | EPI_ISL_3916264 |
| Delta | Delta | AY.44 | EPI_ISL_3916266 |
| Delta | Delta | AY.44 | EPI_ISL_3916267 |

| | | | |
|---|---|---|---|
| Delta | Delta | AY.44 | EPI_ISL_3916268 |
| Delta | Delta | AY.44 | EPI_ISL_3916271 |
| Delta | Delta | AY.44 | EPI_ISL_3916273 |
| Delta | Delta | AY.44 | EPI_ISL_3916274 |
| Delta | Delta | AY.44 | EPI_ISL_3916279 |
| Delta | Delta | AY.44 | EPI_ISL_3916282 |
| Delta | Delta | AY.44 | EPI_ISL_3916285 |
| Delta | Delta | AY.44 | EPI_ISL_3916288 |
| Delta | Delta | AY.44 | EPI_ISL_3916289 |
| Delta | Delta | AY.44 | EPI_ISL_3916291 |
| Delta | Delta | AY.44 | EPI_ISL_3916295 |
| Delta | Delta | AY.44 | EPI_ISL_3916313 |
| Delta | Delta | AY.44 | EPI_ISL_3916317 |
| Delta | Delta | AY.44 | EPI_ISL_3916318 |
| Delta | Delta | AY.44 | EPI_ISL_3916321 |
| Delta | Delta | AY.44 | EPI_ISL_3916329 |
| Delta | Delta | AY.44 | EPI_ISL_3916331 |
| Delta | Delta | AY.44 | EPI_ISL_3916332 |
| Delta | Delta | AY.44 | EPI_ISL_3916337 |
| Delta | Delta | AY.44 | EPI_ISL_3916343 |
| Delta | Delta | AY.44 | EPI_ISL_3916344 |
| Delta | Delta | AY.44 | EPI_ISL_3916346 |
| Delta | Delta | AY.44 | EPI_ISL_3916359 |
| Delta | Delta | AY.44 | EPI_ISL_3916361 |
| Delta | Delta | AY.44 | EPI_ISL_3916363 |
| Delta | Delta | AY.44 | EPI_ISL_3916374 |
| Delta | Delta | AY.44 | EPI_ISL_3916379 |
| Delta | Delta | AY.44 | EPI_ISL_3916381 |
| Delta | Delta | AY.44 | EPI_ISL_3916384 |
| Delta | Delta | AY.44 | EPI_ISL_3916387 |
| Delta | Delta | AY.44 | EPI_ISL_4072015 |
| Delta | Delta | AY.44 | EPI_ISL_4072018 |
| Delta | Delta | AY.46.2 | EPI_ISL_3916303 |
| Delta | Delta | AY.47 | EPI_ISL_3390724 |
| Delta | Delta | AY.47 | EPI_ISL_3722213 |
| Delta | Delta | AY.47 | EPI_ISL_3916265 |
| Delta | Delta | AY.47 | EPI_ISL_3916356 |
| Delta | Delta | AY.47 | EPI_ISL_3916360 |
| Delta | Delta | AY.47 | EPI_ISL_3916378 |
| Delta | Delta | AY.47 | EPI_ISL_3916380 |
| Delta | Delta | AY.47 | EPI_ISL_3916382 |
| Delta | Delta | AY.48 | EPI_ISL_3916306 |
| Delta | Delta | AY.52 | EPI_ISL_3236163 |
| Delta | Delta | AY.54 | EPI_ISL_3390733 |
| Delta | Delta | AY.54 | EPI_ISL_3722178 |
| Delta | Delta | AY.54 | EPI_ISL_3722207 |
| Delta | Delta | AY.59 | EPI_ISL_4496847 |
| Delta | Delta | AY.62 | EPI_ISL_3236152 |
| Delta | Delta | AY.67 | EPI_ISL_3538752 |
| Delta | Delta | AY.67 | EPI_ISL_3916292 |
| Delta | Delta | AY.67 | EPI_ISL_3916293 |
| Delta | Delta | AY.74 | EPI_ISL_3916281 |
| Delta | Delta | AY.75 | EPI_ISL_3050203 |
| Delta | Delta | AY.75 | EPI_ISL_3236150 |
| Delta | Delta | AY.75 | EPI_ISL_3236151 |
| Delta | Delta | AY.75 | EPI_ISL_3236168 |

| | | | |
|---|---|---|---|
| Delta | Delta | AY.75 | EPI_ISL_3236170 |
| Delta | Delta | AY.75 | EPI_ISL_3236182 |
| Delta | Delta | AY.75 | EPI_ISL_3390738 |
| Delta | Delta | AY.75 | EPI_ISL_3538742 |
| Delta | Delta | AY.75 | EPI_ISL_3916284 |
| Delta | Delta | AY.75 | EPI_ISL_4072019 |
| Delta | Delta | AY.98.1 | EPI_ISL_3916392 |
| Delta | Delta | B.1.617.2 | EPI_ISL_2457061 |
| Delta | Delta | B.1.617.2 | EPI_ISL_3050180 |
| Delta | Delta | B.1.617.2 | EPI_ISL_3050191 |
| Delta | Delta | B.1.617.2 | EPI_ISL_3050204 |
| Delta | Delta | B.1.617.2 | EPI_ISL_3236139 |
| Delta | Delta | B.1.617.2 | EPI_ISL_3236143 |
| Delta | Delta | B.1.617.2 | EPI_ISL_3722173 |
| Delta | Delta | B.1.617.2 | EPI_ISL_3722176 |
| Delta | Delta | B.1.617.2 | EPI_ISL_3916277 |
| Delta | Delta | B.1.617.2 | EPI_ISL_3916307 |
| Delta | Delta | B.1.617.2 | EPI_ISL_3916326 |
| Delta | Delta | B.1.617.2 | EPI_ISL_3916355 |
| Delta | Delta | B.1.617.2 | EPI_ISL_4072013 |
| Delta | Delta | B.1.617.2 | EPI_ISL_4072020 |
| Delta | Delta | B.1.617.2 | EPI_ISL_4072021 |
| Delta | Delta | B.1.617.2 | EPI_ISL_4072022 |
| Delta | Delta | B.1.617.2 | EPI_ISL_4072023 |
| Delta | Delta | B.1.617.2 | EPI_ISL_4496821 |
| Delta | Delta | B.1.617.2 | EPI_ISL_4496831 |
| Delta | Delta | B.1.617.2 | EPI_ISL_4496832 |
| Delta | Delta | B.1.617.2 | EPI_ISL_4496833 |
| Delta | Delta | B.1.617.2 | EPI_ISL_4496846 |
| Delta | Delta | B.1.617.2 | EPI_ISL_4496855 |
| Delta | Delta | B.1.617.2 | EPI_ISL_4496874 |
| Delta | Delta | B.1.617.2 | EPI_ISL_4496880 |
| Delta | Delta | B.1.617.2 | EPI_ISL_4496888 |
| Delta | Delta | B.1.617.2 | EPI_ISL_4496890 |
| Delta | Delta | B.1.617.2 | EPI_ISL_4496891 |
| Delta | Delta | B.1.617.2 | EPI_ISL_4496892 |
| Gamma | Not a VOC | B.1.621 | EPI_ISL_3236157 |
| Gamma | Not a VOC | B.1.621 | EPI_ISL_4496900 |
| Gamma | Not a VOC | BB.2 | EPI_ISL_3390723 |
| Gamma | Not a VOC | BB.2 | EPI_ISL_3390757 |
| Gamma | Gamma | P.1 | EPI_ISL_3050186 |
| Gamma | Gamma | P.1 | EPI_ISL_3050210 |
| Gamma | Gamma | P.1 | EPI_ISL_3538746 |
| Gamma | Gamma | P.1 | EPI_ISL_3916315 |
| Gamma | Gamma | P.1 | EPI_ISL_4072051 |
| Gamma | Gamma | P.1 | EPI_ISL_4072052 |
| Gamma | Gamma | P.1 | EPI_ISL_4072073 |
| Gamma | Gamma | P.1 | EPI_ISL_4496811 |
| Gamma | Gamma | P.1 | EPI_ISL_4496812 |
| Gamma | Gamma | P.1 | EPI_ISL_4496813 |
| Gamma | Gamma | P.1 | EPI_ISL_4496815 |
| Gamma | Gamma | P.1 | EPI_ISL_4496816 |
| Gamma | Gamma | P.1 | EPI_ISL_4496875 |
| Gamma | Gamma | P.1.10 | EPI_ISL_3050249 |
| Gamma | Gamma | P.1.10 | EPI_ISL_4496856 |
| Gamma | Gamma | P.1.10 | EPI_ISL_4496866 |

| | | | |
|---|---|---|---|
| Gamma | Gamma | P.1.10 | EPI_ISL_4496871 |
| Gamma | Gamma | P.1.10 | EPI_ISL_4496883 |
| Gamma | Gamma | P.1.17 | EPI_ISL_4496877 |
| Gamma | Gamma | P.1.17 | EPI_ISL_4496901 |
| Not a VOC | Not a VOC | A.2.5 | EPI_ISL_3050209 |
| Not a VOC | Not a VOC | A.2.5 | EPI_ISL_3050212 |
| Not a VOC | Not a VOC | A.2.5 | EPI_ISL_3050213 |
| Not a VOC | Not a VOC | A.2.5 | EPI_ISL_3050214 |
| Not a VOC | Not a VOC | A.2.5 | EPI_ISL_3050251 |
| Not a VOC | Not a VOC | A.2.5 | EPI_ISL_4496879 |
| Not a VOC | Not a VOC | B.1 | EPI_ISL_3050206 |
| Not a VOC | Not a VOC | B.1 | EPI_ISL_3722198 |
| Not a VOC | Not a VOC | B.1 | EPI_ISL_4496826 |
| Not a VOC | Not a VOC | B.1.1.318 | EPI_ISL_4496870 |
| Not a VOC | Not a VOC | B.1.1.519 | EPI_ISL_4496824 |
| Not a VOC | Not a VOC | B.1.311 | EPI_ISL_4072038 |
| Not a VOC | Not a VOC | B.1.427 | EPI_ISL_4496835 |
| Not a VOC | Not a VOC | B.1.427 | EPI_ISL_4496836 |
| Not a VOC | Not a VOC | B.1.427 | EPI_ISL_4496889 |
| Not a VOC | Not a VOC | B.1.429 | EPI_ISL_4496817 |
| Not a VOC | Not a VOC | B.1.429 | EPI_ISL_4496818 |
| Not a VOC | Not a VOC | B.1.429 | EPI_ISL_4496819 |
| Not a VOC | Not a VOC | B.1.429 | EPI_ISL_4496822 |
| Not a VOC | Not a VOC | B.1.429 | EPI_ISL_4496823 |
| Not a VOC | Not a VOC | B.1.429 | EPI_ISL_4496834 |
| Not a VOC | Not a VOC | B.1.429 | EPI_ISL_4496837 |
| Not a VOC | Not a VOC | B.1.429 | EPI_ISL_4496845 |
| Not a VOC | Not a VOC | B.1.526 | EPI_ISL_4496810 |
| Not a VOC | Not a VOC | B.1.526 | EPI_ISL_4496814 |
| Not a VOC | Not a VOC | B.1.526 | EPI_ISL_4496825 |
| Not a VOC | Not a VOC | B.1.526 | EPI_ISL_4496827 |
| Not a VOC | Not a VOC | B.1.526 | EPI_ISL_4496872 |
| Not a VOC | Not a VOC | B.1.526 | EPI_ISL_4496884 |
| Not a VOC | Not a VOC | B.1.526 | EPI_ISL_4496894 |
| Not a VOC | Not a VOC | B.1.526 | EPI_ISL_4496899 |
| Not a VOC | Not a VOC | B.1.526 | EPI_ISL_4496902 |
| Not a VOC | Not a VOC | B.1.526 | EPI_ISL_4496903 |
| Not a VOC | Not a VOC | B.1.627 | EPI_ISL_4496862 |
| Not a VOC | Not a VOC | B.1.637 | EPI_ISL_3050248 |
| Not a VOC | Not a VOC | B.1.637 | EPI_ISL_3236140 |
| Not a VOC | Not a VOC | B.1.637 | EPI_ISL_3390720 |
| Not a VOC | Not a VOC | B.1.637 | EPI_ISL_3390721 |
| Not a VOC | Not a VOC | B.1.637 | EPI_ISL_3390722 |
| Not a VOC | Not a VOC | B.1.637 | EPI_ISL_4496852 |
| Not a VOC | Not a VOC | B.1.637 | EPI_ISL_4496854 |
| Not a VOC | Not a VOC | B.1.637 | EPI_ISL_4496878 |
| Not a VOC | Not a VOC | B.1.637 | EPI_ISL_4496886 |
| Not a VOC | Not a VOC | B.1.637 | EPI_ISL_4496887 |
| Not a VOC | Not a VOC | B.1.637 | EPI_ISL_4496895 |
| Not a VOC | Not a VOC | XB | EPI_ISL_3050187 |
| Not a VOC | Not a VOC | XB | EPI_ISL_3050197 |
| Not a VOC | Not a VOC | XB | EPI_ISL_4072042 |
| Not a VOC | Not a VOC | XB | EPI_ISL_4072043 |
| Not a VOC | Not a VOC | XB | EPI_ISL_4496863 |
| Not a VOC | Not a VOC | XB | EPI_ISL_4496864 |
| Not a VOC | Not a VOC | XB | EPI_ISL_4496873 |

| | | | |
|---|---|---|---|
| Not a VOC | Not a VOC | XB | EPI_ISL_4496893 |
| Not a VOC | Not a VOC | XB | EPI_ISL_4496904 |
| Omicron | Omicron | BA.1 | EPI_ISL_7808007 |
| Omicron | Omicron | BA.1 | EPI_ISL_7808009 |
| Omicron | Omicron | BA.1 | EPI_ISL_7808012 |
| Omicron | Omicron | BA.1 | EPI_ISL_8131086 |
| Omicron | Omicron | BA.1 | EPI_ISL_8131087 |
| Omicron | Omicron | BA.1 | EPI_ISL_8131088 |
| Omicron | Omicron | BA.1 | EPI_ISL_8131090 |
| Omicron | Omicron | BA.1 | EPI_ISL_8131091 |
| Omicron | Omicron | BA.1 | EPI_ISL_8131093 |
| Omicron | Omicron | BA.1 | EPI_ISL_8131094 |
| Omicron | Omicron | BA.1 | EPI_ISL_8131096 |
| Omicron | Omicron | BA.1 | EPI_ISL_8131099 |
| Omicron | Omicron | BA.1 | EPI_ISL_8131102 |
| Omicron | Omicron | BA.1 | EPI_ISL_8131103 |
| Omicron | Omicron | BA.1 | EPI_ISL_8131109 |
| Omicron | Omicron | BA.1 | EPI_ISL_8131110 |
| Omicron | Omicron | BA.1 | EPI_ISL_8131111 |
| Omicron | Omicron | BA.1 | EPI_ISL_8131112 |
| Omicron | Omicron | BA.1 | EPI_ISL_8131114 |
| Omicron | Omicron | BA.1 | EPI_ISL_8131116 |
| Omicron | Omicron | BA.1 | EPI_ISL_8131117 |
| Omicron | Omicron | BA.1 | EPI_ISL_8131118 |
| Omicron | Omicron | BA.1 | EPI_ISL_8131119 |
| Omicron | Omicron | BA.1 | EPI_ISL_8131120 |
| Omicron | Omicron | BA.1 | EPI_ISL_8131121 |
| Omicron | Omicron | BA.1 | EPI_ISL_8131122 |
| Omicron | Omicron | BA.1 | EPI_ISL_8131123 |
| Omicron | Omicron | BA.1 | EPI_ISL_8131125 |
| Omicron | Omicron | BA.1 | EPI_ISL_8131128 |
| Omicron | Omicron | BA.1 | EPI_ISL_8406558 |
| Omicron | Omicron | BA.1 | EPI_ISL_8406559 |
| Omicron | Omicron | BA.1 | EPI_ISL_8406561 |
| Omicron | Omicron | BA.1 | EPI_ISL_8406562 |
| Omicron | Omicron | BA.1 | EPI_ISL_8406565 |
| Omicron | Omicron | BA.1 | EPI_ISL_8406570 |
| Omicron | Omicron | BA.1 | EPI_ISL_8406571 |
| Omicron | Omicron | BA.1 | EPI_ISL_8406573 |
| Omicron | Omicron | BA.1 | EPI_ISL_8406574 |
| Omicron | Omicron | BA.1 | EPI_ISL_8406575 |
| Omicron | Omicron | BA.1 | EPI_ISL_8406576 |
| Omicron | Omicron | BA.1 | EPI_ISL_8406579 |
| Omicron | Omicron | BA.1 | EPI_ISL_8406581 |
| Omicron | Omicron | BA.1 | EPI_ISL_8406587 |
| Omicron | Omicron | BA.1 | EPI_ISL_8406590 |
| Omicron | Omicron | BA.1 | EPI_ISL_8406591 |
| Omicron | Omicron | BA.1 | EPI_ISL_8406592 |
| Omicron | Omicron | BA.1 | EPI_ISL_8406593 |
| Omicron | Omicron | BA.1 | EPI_ISL_8406594 |
| Omicron | Omicron | BA.1 | EPI_ISL_8406595 |
| Omicron | Omicron | BA.1 | EPI_ISL_8406598 |
| Omicron | Omicron | BA.1 | EPI_ISL_8435104 |
| Omicron | Omicron | BA.1 | EPI_ISL_8749105 |
| Omicron | Omicron | BA.1 | EPI_ISL_8749114 |
| Omicron | Omicron | BA.1 | EPI_ISL_8749122 |

| | | | |
|---|---|---|---|
| Omicron | Omicron | BA.1 | EPI_ISL_8749124 |
| Omicron | Omicron | BA.1 | EPI_ISL_8749126 |
| Omicron | Omicron | BA.1 | EPI_ISL_8749127 |
| Omicron | Omicron | BA.1 | EPI_ISL_8749129 |
| Omicron | Omicron | BA.1 | EPI_ISL_8749130 |
| Omicron | Omicron | BA.1 | EPI_ISL_8749132 |
| Omicron | Omicron | BA.1 | EPI_ISL_8749133 |
| Omicron | Omicron | BA.1 | EPI_ISL_8749134 |
| Omicron | Omicron | BA.1 | EPI_ISL_8749135 |
| Omicron | Omicron | BA.1 | EPI_ISL_8749136 |
| Omicron | Omicron | BA.1 | EPI_ISL_8749137 |
| Omicron | Omicron | BA.1 | EPI_ISL_8749138 |
| Omicron | Omicron | BA.1 | EPI_ISL_8749139 |
| Omicron | Omicron | BA.1 | EPI_ISL_8749141 |
| Omicron | Omicron | BA.1 | EPI_ISL_8749142 |
| Omicron | Omicron | BA.1 | EPI_ISL_8749143 |
| Omicron | Omicron | BA.1 | EPI_ISL_8749147 |
| Omicron | Omicron | BA.1 | EPI_ISL_8749150 |
| Omicron | Omicron | BA.1 | EPI_ISL_8749151 |
| Omicron | Omicron | BA.1 | EPI_ISL_8749152 |
| Omicron | Omicron | BA.1 | EPI_ISL_8749153 |
| Omicron | Omicron | BA.1 | EPI_ISL_8749155 |
| Omicron | Omicron | BA.1 | EPI_ISL_8749161 |
| Omicron | Omicron | BA.1 | EPI_ISL_8749164 |
| Omicron | Omicron | BA.1 | EPI_ISL_8953837 |
| Omicron | Omicron | BA.1 | EPI_ISL_8953838 |
| Omicron | Omicron | BA.1 | EPI_ISL_8953861 |
| Omicron | Omicron | BA.1 | EPI_ISL_8953864 |
| Omicron | Omicron | BA.1 | EPI_ISL_8953865 |
| Omicron | Omicron | BA.1 | EPI_ISL_8953868 |
| Omicron | Omicron | BA.1 | EPI_ISL_8953869 |
| Omicron | Omicron | BA.1 | EPI_ISL_8953872 |
| Omicron | Omicron | BA.1 | EPI_ISL_8953876 |
| Omicron | Omicron | BA.1 | EPI_ISL_8953877 |
| Omicron | Omicron | BA.1 | EPI_ISL_8953881 |
| Omicron | Omicron | BA.1 | EPI_ISL_8953886 |
| Omicron | Omicron | BA.1 | EPI_ISL_8953890 |
| Omicron | Omicron | BA.1 | EPI_ISL_8953891 |
| Omicron | Omicron | BA.1 | EPI_ISL_8953894 |
| Omicron | Omicron | BA.1 | EPI_ISL_8953898 |
| Omicron | Omicron | BA.1 | EPI_ISL_8953899 |
| Omicron | Omicron | BA.1 | EPI_ISL_8953902 |
| Omicron | Omicron | BA.1 | EPI_ISL_8953908 |
| Omicron | Omicron | BA.1 | EPI_ISL_8953909 |
| Omicron | Omicron | BA.1 | EPI_ISL_8953910 |
| Omicron | Omicron | BA.1 | EPI_ISL_8953912 |
| Omicron | Omicron | BA.1 | EPI_ISL_8953913 |
| Omicron | Omicron | BA.1 | EPI_ISL_8953917 |
| Omicron | Omicron | BA.1 | EPI_ISL_8953918 |
| Omicron | Omicron | BA.1 | EPI_ISL_8953921 |
| Omicron | Omicron | BA.1 | EPI_ISL_8953923 |
| Omicron | Omicron | BA.1 | EPI_ISL_8953924 |
| Omicron | Omicron | BA.1 | EPI_ISL_8953926 |
| Omicron | Omicron | BA.1 | EPI_ISL_8953930 |
| Omicron | Omicron | BA.1 | EPI_ISL_8953932 |
| Omicron | Omicron | BA.1 | EPI_ISL_8953933 |

| | | | |
|---|---|---|---|
| Omicron | Omicron | BA.1 | EPI_ISL_8953934 |
| Omicron | Omicron | BA.1 | EPI_ISL_8953935 |
| Omicron | Omicron | BA.1 | EPI_ISL_8953937 |
| Omicron | Omicron | BA.1 | EPI_ISL_8953938 |
| Omicron | Omicron | BA.1 | EPI_ISL_8953940 |
| Omicron | Omicron | BA.1 | EPI_ISL_8953942 |
| Omicron | Omicron | BA.1 | EPI_ISL_8953949 |
| Omicron | Omicron | BA.1 | EPI_ISL_8953951 |
| Omicron | Omicron | BA.1 | EPI_ISL_8953955 |
| Omicron | Omicron | BA.1 | EPI_ISL_8953957 |
| Omicron | Omicron | BA.1 | EPI_ISL_8953959 |
| Omicron | Omicron | BA.1 | EPI_ISL_8953960 |
| Omicron | Omicron | BA.1 | EPI_ISL_8953961 |
| Omicron | Omicron | BA.1.1 | EPI_ISL_8131089 |
| Omicron | Omicron | BA.1.1 | EPI_ISL_8131092 |
| Omicron | Omicron | BA.1.1 | EPI_ISL_8131095 |
| Omicron | Omicron | BA.1.1 | EPI_ISL_8131097 |
| Omicron | Omicron | BA.1.1 | EPI_ISL_8131098 |
| Omicron | Omicron | BA.1.1 | EPI_ISL_8131100 |
| Omicron | Omicron | BA.1.1 | EPI_ISL_8131101 |
| Omicron | Omicron | BA.1.1 | EPI_ISL_8131104 |
| Omicron | Omicron | BA.1.1 | EPI_ISL_8131105 |
| Omicron | Omicron | BA.1.1 | EPI_ISL_8131106 |
| Omicron | Omicron | BA.1.1 | EPI_ISL_8131107 |
| Omicron | Omicron | BA.1.1 | EPI_ISL_8131108 |
| Omicron | Omicron | BA.1.1 | EPI_ISL_8131113 |
| Omicron | Omicron | BA.1.1 | EPI_ISL_8131115 |
| Omicron | Omicron | BA.1.1 | EPI_ISL_8131124 |
| Omicron | Omicron | BA.1.1 | EPI_ISL_8131126 |
| Omicron | Omicron | BA.1.1 | EPI_ISL_8131127 |
| Omicron | Omicron | BA.1.1 | EPI_ISL_8406560 |
| Omicron | Omicron | BA.1.1 | EPI_ISL_8406563 |
| Omicron | Omicron | BA.1.1 | EPI_ISL_8406564 |
| Omicron | Omicron | BA.1.1 | EPI_ISL_8406566 |
| Omicron | Omicron | BA.1.1 | EPI_ISL_8406567 |
| Omicron | Omicron | BA.1.1 | EPI_ISL_8406569 |
| Omicron | Omicron | BA.1.1 | EPI_ISL_8406572 |
| Omicron | Omicron | BA.1.1 | EPI_ISL_8406577 |
| Omicron | Omicron | BA.1.1 | EPI_ISL_8406580 |
| Omicron | Omicron | BA.1.1 | EPI_ISL_8406582 |
| Omicron | Omicron | BA.1.1 | EPI_ISL_8406583 |
| Omicron | Omicron | BA.1.1 | EPI_ISL_8406584 |
| Omicron | Omicron | BA.1.1 | EPI_ISL_8406585 |
| Omicron | Omicron | BA.1.1 | EPI_ISL_8406586 |
| Omicron | Omicron | BA.1.1 | EPI_ISL_8406588 |
| Omicron | Omicron | BA.1.1 | EPI_ISL_8406589 |
| Omicron | Omicron | BA.1.1 | EPI_ISL_8406596 |
| Omicron | Omicron | BA.1.1 | EPI_ISL_8406597 |
| Omicron | Omicron | BA.1.1 | EPI_ISL_8749096 |
| Omicron | Omicron | BA.1.1 | EPI_ISL_8749123 |
| Omicron | Omicron | BA.1.1 | EPI_ISL_8749125 |
| Omicron | Omicron | BA.1.1 | EPI_ISL_8749128 |
| Omicron | Omicron | BA.1.1 | EPI_ISL_8749131 |
| Omicron | Omicron | BA.1.1 | EPI_ISL_8749140 |
| Omicron | Omicron | BA.1.1 | EPI_ISL_8749144 |
| Omicron | Omicron | BA.1.1 | EPI_ISL_8749145 |

| | | | |
|---|---|---|---|
| Omicron | Omicron | BA.1.1 | EPI_ISL_8749146 |
| Omicron | Omicron | BA.1.1 | EPI_ISL_8749148 |
| Omicron | Omicron | BA.1.1 | EPI_ISL_8749149 |
| Omicron | Omicron | BA.1.1 | EPI_ISL_8749154 |
| Omicron | Omicron | BA.1.1 | EPI_ISL_8749156 |
| Omicron | Omicron | BA.1.1 | EPI_ISL_8749157 |
| Omicron | Omicron | BA.1.1 | EPI_ISL_8749158 |
| Omicron | Omicron | BA.1.1 | EPI_ISL_8749159 |
| Omicron | Omicron | BA.1.1 | EPI_ISL_8749160 |
| Omicron | Omicron | BA.1.1 | EPI_ISL_8749162 |
| Omicron | Omicron | BA.1.1 | EPI_ISL_8749163 |
| Omicron | Omicron | BA.1.1 | EPI_ISL_8749165 |
| Omicron | Omicron | BA.1.1 | EPI_ISL_8953860 |
| Omicron | Omicron | BA.1.1 | EPI_ISL_8953862 |
| Omicron | Omicron | BA.1.1 | EPI_ISL_8953863 |
| Omicron | Omicron | BA.1.1 | EPI_ISL_8953866 |
| Omicron | Omicron | BA.1.1 | EPI_ISL_8953867 |
| Omicron | Omicron | BA.1.1 | EPI_ISL_8953870 |
| Omicron | Omicron | BA.1.1 | EPI_ISL_8953871 |
| Omicron | Omicron | BA.1.1 | EPI_ISL_8953873 |
| Omicron | Omicron | BA.1.1 | EPI_ISL_8953874 |
| Omicron | Omicron | BA.1.1 | EPI_ISL_8953875 |
| Omicron | Omicron | BA.1.1 | EPI_ISL_8953878 |
| Omicron | Omicron | BA.1.1 | EPI_ISL_8953879 |
| Omicron | Omicron | BA.1.1 | EPI_ISL_8953880 |
| Omicron | Omicron | BA.1.1 | EPI_ISL_8953882 |
| Omicron | Omicron | BA.1.1 | EPI_ISL_8953883 |
| Omicron | Omicron | BA.1.1 | EPI_ISL_8953884 |
| Omicron | Omicron | BA.1.1 | EPI_ISL_8953885 |
| Omicron | Omicron | BA.1.1 | EPI_ISL_8953887 |
| Omicron | Omicron | BA.1.1 | EPI_ISL_8953888 |
| Omicron | Omicron | BA.1.1 | EPI_ISL_8953889 |
| Omicron | Omicron | BA.1.1 | EPI_ISL_8953892 |
| Omicron | Omicron | BA.1.1 | EPI_ISL_8953893 |
| Omicron | Omicron | BA.1.1 | EPI_ISL_8953895 |
| Omicron | Omicron | BA.1.1 | EPI_ISL_8953900 |
| Omicron | Omicron | BA.1.1 | EPI_ISL_8953901 |
| Omicron | Omicron | BA.1.1 | EPI_ISL_8953903 |
| Omicron | Omicron | BA.1.1 | EPI_ISL_8953904 |
| Omicron | Omicron | BA.1.1 | EPI_ISL_8953905 |
| Omicron | Omicron | BA.1.1 | EPI_ISL_8953906 |
| Omicron | Omicron | BA.1.1 | EPI_ISL_8953907 |
| Omicron | Omicron | BA.1.1 | EPI_ISL_8953911 |
| Omicron | Omicron | BA.1.1 | EPI_ISL_8953914 |
| Omicron | Omicron | BA.1.1 | EPI_ISL_8953915 |
| Omicron | Omicron | BA.1.1 | EPI_ISL_8953916 |
| Omicron | Omicron | BA.1.1 | EPI_ISL_8953919 |
| Omicron | Omicron | BA.1.1 | EPI_ISL_8953920 |
| Omicron | Omicron | BA.1.1 | EPI_ISL_8953922 |
| Omicron | Omicron | BA.1.1 | EPI_ISL_8953925 |
| Omicron | Omicron | BA.1.1 | EPI_ISL_8953927 |
| Omicron | Omicron | BA.1.1 | EPI_ISL_8953928 |
| Omicron | Omicron | BA.1.1 | EPI_ISL_8953929 |
| Omicron | Omicron | BA.1.1 | EPI_ISL_8953931 |
| Omicron | Omicron | BA.1.1 | EPI_ISL_8953936 |
| Omicron | Omicron | BA.1.1 | EPI_ISL_8953939 |

| | | | |
|---|---|---|---|
| Omicron | Omicron | BA.1.1 | EPI_ISL_8953941 |
| Omicron | Omicron | BA.1.1 | EPI_ISL_8953950 |
| Omicron | Omicron | BA.1.1 | EPI_ISL_8953952 |
| Omicron | Omicron | BA.1.1 | EPI_ISL_8953953 |
| Omicron | Omicron | BA.1.1 | EPI_ISL_8953954 |
| Omicron | Omicron | BA.1.1 | EPI_ISL_8953956 |
| Omicron | Omicron | BA.1.1 | EPI_ISL_8953958 |
| Omicron | Omicron | BA.1.1 | EPI_ISL_8953962 |

VOC, variant of concern

[a] Pangolin version 3.1.17

**Supplemental Table 4.** Patient Characteristics and Aggregate Genotyping RT-qPCR Screening Results During Initial Study Period (n=1,657)

| Characteristic | All SARS-CoV-2 Infections | RT-qPCR Subset | WGS Validation Subset |
|---|---|---|---|
| Patients | 1,657 | 1,093 | 502 |
| Age at First Positive NAAT | 33 [21-49] | 33 [22-49] | 33 [22-48] |
| Female | 842 (51%) | 571 (52%) | 265 (53%) |
| Initial Diagnostic Specimen | - | - | - |
| Month of Diagnosis | - | - | - |
| May 2021 & prior | 416 (25%) | 299 (27%) | 87 (17%) |
| June 2021 | 224 (14%) | 149 (14%) | 77 (15%) |
| July 2021 & after | 1017 (61%) | 645 (59%) | 338 (67%) |
| Specimen Collection Site | - | - | - |
| Outpatient COVID-19 Screening Site | 658 (40%) | 412 (38%) | 198 (39%) |
| Emergency Department | 392 (24%) | 287 (26%) | 104 (21%) |
| Outpatient Laboratory | 167 (10%) | 100 (9%) | 57 (11%) |
| Occupational/Student Health | 160 (10%) | 104 (10%) | 67 (13%) |
| Outpatient Clinic | 105 (6%) | 57 (5%) | 36 (7%) |
| Urgent Care | 80 (5%) | 63 (6%) | 34 (7%) |
| Inpatient | 49 (3%) | 30 (3%) | 4 (1%) |
| Perioperative/Periprocedural | 46 (3%) | 40 (4%) | 2 (<1%) |
| Specimen Source | - | - | - |
| Nasopharyngeal Swab | 989 (59%) | 661 (61%) | 281 (56%) |
| Mid-Turbinate Nasal Swab | 659 (40%) | 428 (39%) | 219 (44%) |
| Other[a] | 9 (1%) | 4 (<1%) | 2 (<1%) |
| NAAT Platform | - | - | - |
| Hologic Panther[b] | 869 (52%) | 512 (47%) | 246 (49%) |
| Cepheid GeneXpert[c] | 234 (14%) | 180 (16%) | 87 (17%) |
| Applied Biosystems QuantStudio[b] | 220 (13%) | 155 (14%) | 93 (19%) |
| Genmark Eplex[c] | 169 (10%) | 133 (12%) | 19 (4%) |
| Qiagen Rotor-Gene LDT[b] | 136 (8%) | 85 (8%) | 44 (9%) |
| Roche Liat[c] | 29 (2%) | 28 (3%) | 13 (3%) |
| Variant of Concern | - | - | - |
| Alpha | - | 150 (14%) | 43 (9%) |
| Beta[d] | - | 6 (1%) | 2 (<1%) |
| Gamma[d] | - | 32 (3%) | 20 (4%) |
| Delta | - | 660 (60%) | 378 (75%) |
| Not a Variant of Concern | - | 93 (9%) | 59 (12%) |
| Unable to Genotype[e] | - | 152 (14%) | - |

Data provided as N (column %) or median [interquartile range]
NAAT, nucleic acid amplification test; LDT, lab-developed test; RT-qPCR, reverse transcription quantitative polymerase chain reaction; WGS, whole-genome sequencing
[a] Other includes Nasal Swab, Lung BAL, Oropharynx
[b] Laboratory-based testing methods
[c] Near-care testing methods
[d] RT-qPCR cannot distinguish variants of concern Beta and Gamma variants from variant of interest Mu.
[e] Low viral load, amplification failure, or reaction set-up failure.
